# OMKar automates genome karyotyping using optical maps to identify constitutional abnormalities

**DOI:** 10.1101/2025.02.13.25322211

**Authors:** Siavash Raeisi Dehkordi, Zhaoyang Jia, Joey Estabrook, Jen Hauenstein, Neil Miller, Naz Güleray-Lafci, Jürgen Neesen, Alex Hastie, Alka Chaubey, Andy Wing Chun Pang, Paul Dremsek, Vineet Bafna

**Affiliations:** Department of Computer Science & Engineering, UC San Diego, La Jolla, California, USA; Bionano Genomics, Inc., 9540 Towne Centre Drive Suite 100, San Diego, CA 92121; Halicioğlu Data Science Institute, UC San Diego, La Jolla, California, USA; Institute of Medical Genetics, Center for Pathobiochemistry and Genetics, Medical University of Vienna, 1090 Vienna, Austria

## Abstract

The whole-genome karyotype refers to the sequence of large chromosomal segments comprising an individual’s genotype. Karyotype analysis, which includes identifying aneuploidies and structural rearrangements, is essential for understanding genetic risk factors, informing diagnosis and treatment, and guiding genetic counseling in constitutional disorders. The current karyotyping standard relies on microscopic chromosome examination, a complex and expertise dependent process with megabase scale resolution.

Optical Genome Mapping (OGM) technology offers an efficient approach to detect large-scale genomic lesions. Here, we introduce OMKar, a computational method that generates virtual karyotypes from OGM data. OMKar integrates structural variants (SVs) and copy number (CN) variants into a breakpoint graph representation. It re-estimates copy numbers using Integer Linear Programming to enforce CN balance, and then identifies constrained Eulerian paths corresponding to full chromosome structures.

OMKar was evaluated on 38 whole-genome simulations of constitutional disorders, achieving 88% precision and 95% recall for SV concordance and a 95% Jaccard score for CN concordance. We further applied OMKar to 154 clinical samples including 50 prenatal, 41 postnatal, and 63 parental genomes collected across ten sites. It correctly reconstructed the karyotype in 144 cases, including 25 of 25 aneuploidies, 32 of 32 balanced translocations, and 72 of 82 unbalanced rearrangements. Identified disorders included Cri-du-chat, Wolf-Hirschhorn, Prader-Willi, Down, and Turner syndromes. Notably, OMKar uncovered plausible genetic mechanisms in five previously unexplained cases. These results demonstrate the accuracy and utility of OMKar for OGM-based constitutional karyotyping.

## Introduction

Genomic structural variants involving the loss, amplification, or rearrangement of large genomic regions have been associated with many constitutional diseases(Stankiewicz and Lupski, 2010). The Decipher database lists over 2500 disorders, often caused by large structural changes in the genome, including trisomy, microdeletions and duplications, and other rearrangements(Firth et al., 2009). For molecular diagnosis, affected individuals undergo standard of care (SOC) testing from drawn blood, where the extracted DNA is analyzed for genetic lesions. Genetic prenatal testing is also an important need, despite the recent advancements of non-invasive screening (NIPS) methods, typically utilizing maternal blood samples. Data from large studies suggest that while the negative predictive value (TN/(FN+TN)) was close to 100% for Down syndrome (trisomy 21) screening, the precision was in the 50-81% range, and the numbers were similar for other disorders(Bianchi et al., 2014; Taylor-Phillips et al., 2016). Thus, a positive NIPS result is typically followed by a more invasive molecular diagnostic.

The current standard of care for genetic diagnostic tests includes (a) karyotyping, (b) chromosomal microarray (CMA)(Miller et al., 2010; Shinawi and Cheung, 2008), (c) FISH screening(Pergament et al., 2000; de Moraex-Malinverni et al., 2016), (d) panel sequencing, or (e) whole exome sequencing. Karyotyping methods require considerable manual expertise and have a low resolution of 3-10 Mbp(Shaffer and Bejjani, 2004). They can be combined with CMA or whole exome sequencing to improve resolution for detecting copy number changes. These high resolution methods (CMA, panel sequencing) do not easily detect copy number neutral rearrangements. FISH requires knowledge of probes and is therefore limited in detecting novel variations. In contrast, about 50% of all reciprocal translocations are *de novo*(Chang et al., 2013). Balanced rearrangements are found in 0.2% of individuals (up to 2.2% of the individuals with a previous history of miscarriage). Individuals with a balanced translocation may not directly present with a phenotype/syndrome, but during meiosis, a gamete could carry an unbalanced copy number and result in fertility issues(Chantot-Bastaraud et al., 2008; Dai et al., 2022). However, balanced translocations would be very likely missed by exome sequencing/CMA.

Optical Genome Mapping (OGM) provides an exciting alternative to diagnostic technologies that lie between cytogenetics and exome sequencing in terms of resolution. OGMs are large enough to span repetitive and low complexity regions, while still being able to capture smaller structural variations. While OGM technology cannot call single nucleotide substitutions, or small insertions and deletions, it is well-suited for calling aneuploidies, larger structural variations, balanced and unbalanced rearrangements, inversions and deletions(Nilius-Eliliwi et al., 2023; Balducci et al., 2022), especially with the development of advanced tools(Raeisi Dehkordi et al., 2021; Li et al., 2017). OGM has been used to successfully identify constitutional genomic lesions, despite some limitations(Mantere et al., 2021; Dai et al., 2022; Dremsek et al., 2021; Sahajpal et al., 2021). In principle, OGMs can be supplanted by long read whole genome sequencing(Goenka et al., 2022), but these methods are not yet readily available in a clinical setting. In fact, the demand for OGM based diagnostics is increasing(Levy et al., 2024; Ghabrial et al., 2024; Iqbal et al., 2023).

### Automated karyotyping using OGM

The molecular karyotype of a donor can be described as a collection of genomic sequences, each sequence corresponding to one donor chromosome. Traditionally, the karyotype information was captured by cytogenetics, albeit at low resolution, and helped identify balanced and unbalanced rearrangements, aneuploidies, and other events that are directly relevant to constitutional disorders. In moving from cytogenetics to CMA and exome sequencing, much of that important information was lost. Current methods for SV calling typically do not capture the larger karyotype making it harder to assign significance, for example, to a translocation event, or determining the locations of amplified genomic segments(Xiao et al., 2024). Here, we present a method, *OMKar*, for automatically identifying karyotypes using OGM data.

We tested our method using extensive simulations as well as on OGM data acquired from over 100 prenatal and postnatal samples with constitutional disorders to gain an improved understanding of the power and limitations of the OGM technology for karyotyping.

## Results

### An Overview of OMKar

A brief outline of OMKar (Fig. 1A-D) is presented here, with details in Methods. OMKar processes the output of the Bionano Solve pipeline(BionanoGenomics, 2018), which includes structural variation (SV), copy number variation (CNV), and contig alignment data. It generates a *molecular karyotype* (Table 1) in a custom text format (Supplementary Section S1) and also presents the karyotype as chromosomal clusters using ISCN language, with reference genome coordinates instead of cytogenetic bands. This approach bridges karyotyping and SV calling. Additionally, OMKar provides a graphical karyotype display.

**Table 1:** Terminology.

|  |  |
| --- | --- |
| **Segment.** | An oriented, continuous genomic interval from the reference genome, denoted by (chromosome, start-coordinate, end-coordinate). A **donor chromosome** is described as an ordered sequence of segments. |
| **Breakpoint.** | A breakpoint is described by a pair of non-adjacent coordinates denoting a transition from one segment to another in the donor. |
| **Chromosome group.** | A set of all homologous donor chromosomes sharing the same chromosomal identity. The chromosomal identity is determined by the most represented centromere, and if the chromosome is acentric, by the most represented chromosomal origin of its composing segments. |
| **Chromosome cluster.** | A pair of chromosome groups is denoted as dependent if there exist breakpoints connecting them. A **chromosome cluster** is a connected component of dependent chromosome groups. A chromosome cluster is often defined by a set of **canonical structural variants**, each with an ISCN nomenclature (International Standard of Cytogenetic Nomenclature). |
| **Molecular karyotype.** | A proposed file format that unambiguously describes the karyotype in terms of segments, with nucleotide-level resolution. This file format contains a dictionary of segments that span the entire reference genome, followed by a set of ordered sequences of segments, each representing a chromosome. |

**Figure 1.**
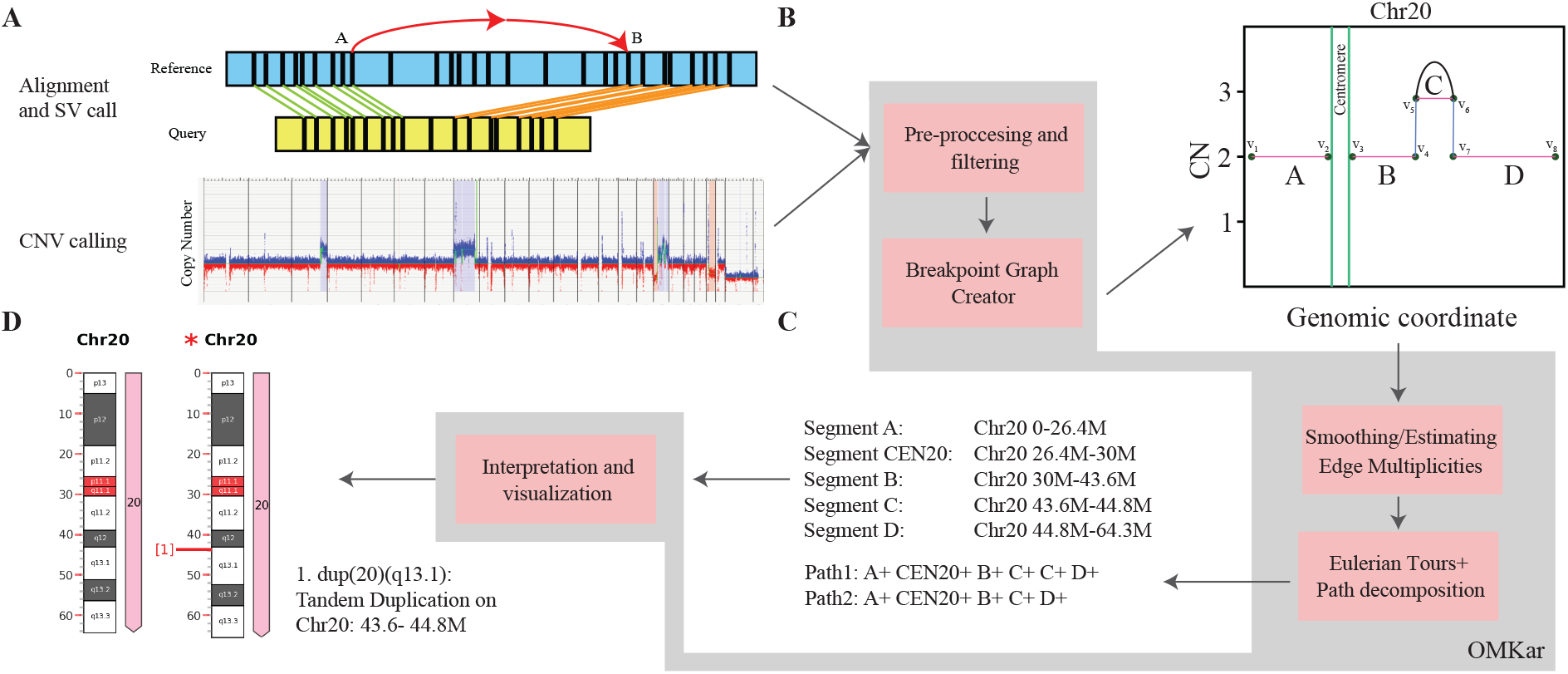
Overview of the OMKar method. (A) Input data: OMKar takes structural variant (SV) calls, copy number variation (CNV) calls, and sequence alignments as input. (B) Pre-processing and breakpoint graph construction after filtering SV and CNV calls to remove low-confidence variations. Chromosomes are segmented based on CNV boundaries and breakpoints, and a breakpoint graph is constructed, where vertices represent segment boundaries and edges represent segment continuity, reference adjacencies, and rearrangements. (C) Smoothing and path decomposition: Integer Linear Programming is used to estimate edge multiplicities, ensuring consistency of copy number constraints. Edge editing is used to generate a Euclidean graph, and paths extraction methods used to reconstruct chromosomes. (D) Interpretation and visualization: Structural variations are re-coded using ISCN, disrupted genes are identified, and results are compiled into an interactive HTML report with chromosome visualizations.

Prior to our work, formal measurements of karyotyping accuracy were lacking. To address this, we developed two additional tools: KarSim, which generates random karyotypes in molecular karyotype and FASTA formats, and KarCheck, which compares two karyotypes by measuring their SV and CNV similarities. These tools help improve method comparisons and enable cross-technology evaluations.

The OMKar algorithm follows a multi-step process: (a) pre-processing of input data, (b) construction of a breakpoint graph, (c) smoothing of edge multiplicities to generate an Eulerian graph, (d) computation of an Eulerian tour, and (e) chromosomal segregation and identification to derive the molecular karyotype. These steps are detailed in Methods.

### OMKar runs efficiently on a desktop server

We tested the tool’s performance on 154 clinical samples and 38 simulated datasets using a standard linux machine (Intel(R) Xeon(R) CPU X5680 @ 3.33GHz, 128 GB of RAM, and running Ubuntu 16.04.6 LTS.) While this represents a relatively powerful configuration, OMKar is designed to be efficient in computational resources, requiring only a single core and with a max RAM usage of 6 GB through all samples tested. OMKar was very efficient with a median runtime of 8.4s (range [6.2-26.1s]; Supplementary Fig. S1). Including the time for image generation required for the html output, the median runtime increased to 21.3s (range 15.0-48.5s). The runtime was correlated with the number of rearrangements (breakpoints).

### OMKar reconstructs karyotypes in simulated data with high accuracy

A total of 552 structural variations were simulated on 38 karyotypes at 100× coverage. The 38 karyotypes could be grouped into 803 chromosome clusters. 299 of 803 clusters had at least one SV. Of the remaining 504 ‘non-event’ clusters, only 6 were reconstructed with an SV (false-positive), yielding a true-negative rate of 98.8%. We observed that the Bionano variant calling pipeline had a lower accuracy of 42.7% in capturing terminal SVs, which were simulated in the peri-telomeric regions (Supplementary Section S2, Supplementary Table S1). Therefore, in the following, we focused on clusters that contain non-terminal SV, represented in 250 of 299 chromosomal clusters.

We first tested if OMKar could estimate the number of chromosomes correctly. 14 aneuploidies– a gain or loss of a chromosome–where simulated in 9 of the 250 chromosome clusters with events, and OMKar correctly reconstructed 13 of them. OMKar reconstructed a normal number of chromosomes in 229 of the remaining 241 clusters (6 FP in the 208 clusters without terminal events, each with three or more balanced translocations; 6 FP in the 33 clusters containing terminal events, due to arm deletion).

For each cluster containing non-terminal SVs, we computed the Jaccard similarity (intersection over union) of the non-terminal SVs in the simulated and predicted clusters. The average Jaccard Similarity across the 250 clusters was 84.8% (recall 94.7%; precision 87.5%), suggesting a highquality karyotype reconstruction. To quantify performance according to the relative difficulty of the simulation, we estimated the *complexity* of each cluster as the number of breakpoint-edges from non-terminal SVs. We denoted clusters with complexity score ≤ 6 as being *low-complexity*, and *high-complexity* otherwise. As expected, the performance on the low-complexity clusters (Fig. 2A; Jaccard 89.9%; Recall 95.6%; Precision 92.2%) was much better than on high-complexity clusters (Jaccard 80.8%; Recall 89.9%, precision 85.8%). The CNV comparison metric behaved similarly with high average Jaccard Similarity across the 250 clusters at 96.0% and some degradation in performance from low to high complexity (Fig. 2B). The clinical cases described in the next section were of lower complexity (Fig. 2C). The distribution of the SV sizes suggests good representation of each SV type (Supplementary Fig. S2).

**Figure 2.**
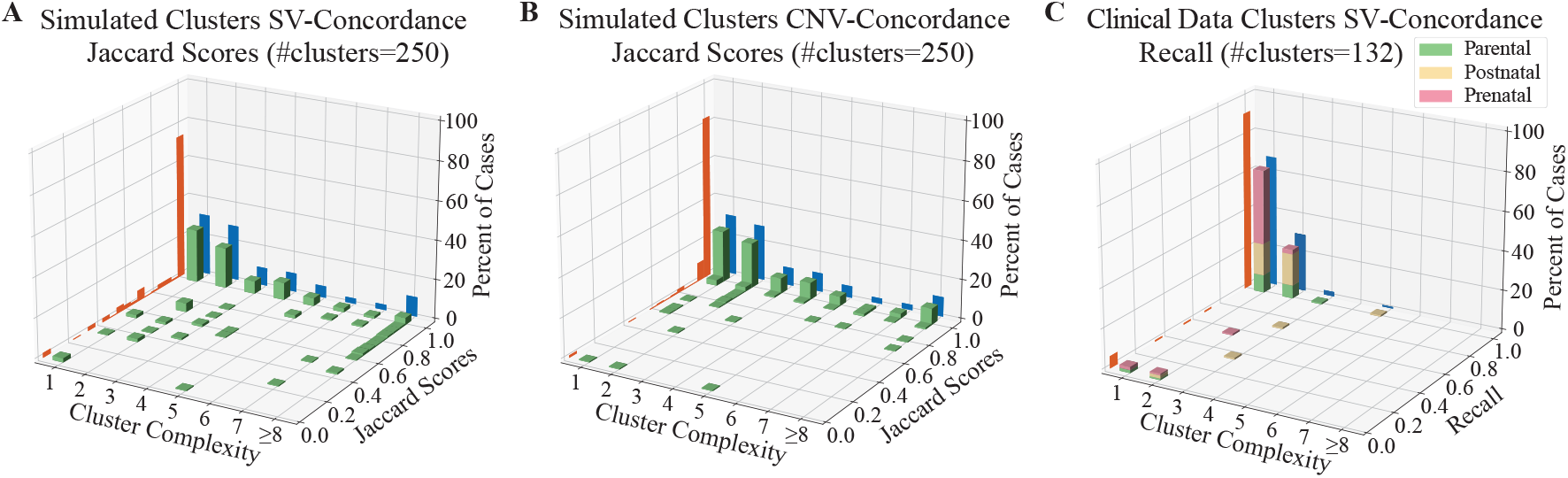
Validation statistics on simulated and clinical karyotypes. Each plot displays a 3-d histogram of Jaccard score (or Recall) of clusters in the sample group. The three axes separate clusters based on complexity, frequency of observations, and Jaccard score or Recall. The frequencies of specific Jaccard scores are displayed in the orange projection, showing that the vast majority of samples have high Jaccard score or Recall. The frequencies of cluster complexity are shown in blue and reveal that the cluster complexity of simulated cases is generally higher than clinical data. (a) Jaccard score of SV edges in simulations; (b) Jaccard score of CNV calls in simulations; (c) Recall of SV calls on 132 clusters from prenatal, postnatal and parental clinical samples.

In addition, we measured accuracy by directly investigating the SV edges (breakpoints) in the chromosome clusters (Table 2). A total of 839 SV edges were introduced in simulations, with 502 in low-complexity clusters. The overall accuracy was high: 96% for low-complexity and 89% for high-complexity clusters, where the occurrence of closely spaced SV edges created challenges. The accuracy also varied depending on the SV type. For example, OMKar was successful in catching balanced translocations but had lower accuracy for Tandem Duplications and Duplication Inversions. These results support the conclusion that OMKar can accurately reconstruct the karyotypes, especially in samples with low rearrangement complexity.

**Table 2:**
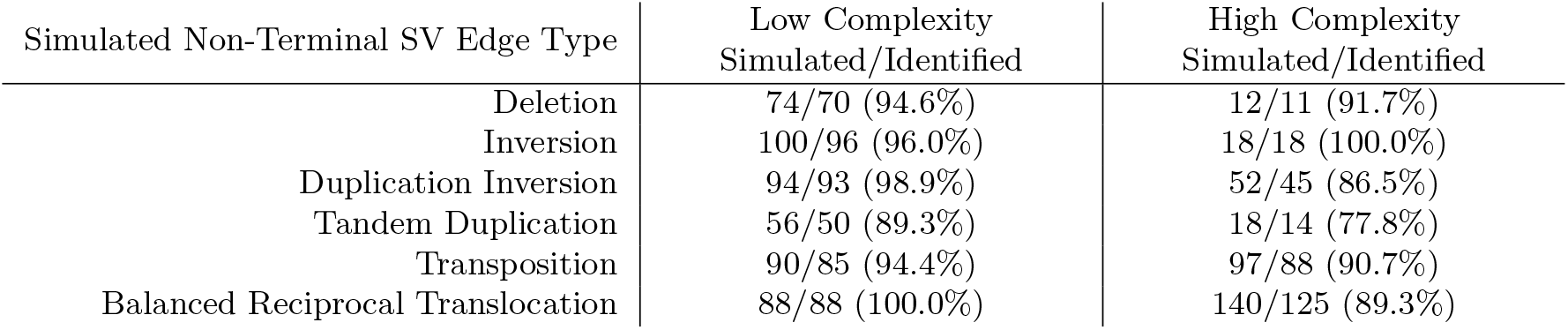
Simulated Non-Terminal SV Edge Recall by Event Types.

| Simulated Non-Terminal SV Edge Type | Low Complexity<br>Simulated/Identified | High Complexity<br>Simulated/Identified |
| --- | --- | --- |
| Deletion | 74/70 (94.6%) | 12/11 (91.7%) |
| Inversion | 100/96 (96.0%) | 18/18 (100.0%) |
| Duplication Inversion | 94/93 (98.9%) | 52/45 (86.5%) |
| Tandem Duplication | 56/50 (89.3%) | 18/14 (77.8%) |
| Transposition | 90/85 (94.4%) | 97/88 (90.7%) |
| Balanced Reciprocal Translocation | 88/88 (100.0%) | 140/125 (89.3%) |

### OMKar reconstructs karyotypes from prenatal, postnatal, and parental screenings with high accuracy

We applied OMKar to OGM data acquired from 154 samples (50 prenatal, 41 postnatal, and 63 parental) prepared at ten different sites (Supplementary Table S2). Seven postnatal samples were biological replicates of 3 individuals, including samples that were mapped at different test sites. For each sample, a previous diagnosis of constitutional abnormality had been made using combinations of traditional cytogenetic methods of karyotyping, CMA, and FISH. These methods are not comprehensive, but they have high precision, so we first checked if OMKar could correctly reconstruct previously detected variations.

The union of calls from karyotyping, CMA, and FISH revealed 141 variations in 154 samples. OMKar was able to fully reconstruct 129 (91%) of the 141 variations including 25/25 (100%) aneuploidies, 32/32 (100%) balanced reciprocal translocations, 38/39 (97.4%) deletions, 32/38 (84.2%) amplifications, 1/3 (33.3%) unbalanced translocations. OMKar did not detect 1 inversion, 1 Robertsonian translocation, and 1 isodicentric chromosome. These 146 structural variations formed 132 chromosome clusters, where reconstructions of 121 (91.7%) were fully concordant. Some of the missed karyotypes were due to SVs being masked (3 clusters), duplication size below OMKar threshold (2 clusters), Robertsonian Translocation (1 cluster), and isodicentric chromosome (1 cluster; Fig. 2C, Supplementary Table S3).

Importantly, OMKar improved upon every other technology when considered in isolation (Supplementary Fig. S3). On the 65 samples where karyotyping was performed, it detected only 56 (64%) of the 87 SVs. Similarly, CMA was applied on 76 samples and detected 94 (88%) of the 107 SVs; FISH was applied on 16 samples, and detected 11 (58%) of the 19 SVs. Specifically, karyotyping mostly captured large rearrangements, whereas CMA and FISH mostly captured unbalanced events.

We tested OMKar consistency using biological replicates prepared and mapped at different test sites. Specifically, one postnatal sample was processed at six different test sites, and two postnatal samples were each processed at two test sites (Supplementary Table S2). In all cases, OMKar successfully reconstructed the correct karyotype.

Importantly, OMKar reconstructed additional SVs not caught by any of the other techniques. Specifically, after filtering out lower quality SVs (Section S3.1), OMKar detected 436 deletions, 506 amplifications and 67 inversions, averaging 2.8 deletions, 3.3 amplifications, and 0.44 inversions as novel events per sample. These discoveries need to be experimentally validated. However, coverage support for novel SVs was similar to simulations, where OMKar achieved a precision of 88%.

### In contrast with Bionano Access, OMKar reconstructs the full karyotypes

OMKar takes as input the SV and CNV calls generated by Bionano Solve, which are also available through the Bionano Access software. In principle, a trained cytogeneticist could manually reconstruct the complete karyotype using Bionano Access information. However, in practice, particularly for samples with complex structural rearrangements, manual reconstruction is time-consuming and has potential for error. OMKar overcomes this limitation by fully automating this process, producing fast and consistent karyotype reconstructions.

To demonstrate the added value of OMKar, we compared its results with those presented by Bionano Access in three postnatal cases involving translocations. These included one canonical balanced reciprocal translocation and two cases with multiple, complex rearrangements.

Postnatal sample 1404 contains a balanced reciprocal translocation between Chromosomes 9p and 22q, along with a 240 kbp left-duplication inversion on Chromosome 22 located approximately 1.3 Mbp upstream of the translocation breakpoint. Bionano Access correctly reported the presence of each individual event as discordant breakpoint edges (Supplementary Fig. S4A). In contrast, OMKar reconstructed the derivative chromosomes in their entirety, resolving the rearranged segment order (e.g., … 74+ 75-75+ 76+ 30-in derivative Chromosome 22; Supplementary Fig. S4B), where each number represents a distinct chromosomal segment and annotates the overall karyotype structure. It also generated an ISCN style representation (Supplementary Fig. S4C), and a chromosome ideogram (Supplementary Fig. S4D). An annotation of the balanced translocations and inverted duplication are provided for clarity (Supplementary Fig. S4E).

Postnatal sample 2281 (Supplementary Fig. S5) contains a set of complex inter-chromosomal rearrangements between the q-arm of Chromosome 2 and the p-arm of Chromosome 3. The Bionano Access output reports four distinct inter-chromosomal translocations, along with one deletion and one inversion. The four translocations share two approximate breakpoints but differ in orientation. Manual reconstruction of the karyotype by a cytogeneticist using Bionano Access information would be highly time-consuming. In contrast, OMKar generated a complete and coherent karyotype reconstruction without adding or omitting any calls (Supplementary Fig. S5B-D).

Postnatal sample 2282 (Supplementary Fig. S6) contains a set of complex inter- and intrachromosomal rearrangements between the p-arms of Chromosome 12 and 16. Bionano Access reports one inter-chromosomal translocation between the two chromosomes, followed by an intrachromosomal translocation, a deletion, and an inversion downstream of the initial translocation breakpoint on Chromosome 16. A copy number gain is also observed on Chromosome 16 near the intra-chromosomal translocation breakpoint. Notably, the CN gain does not span the entire region between the translocation breakpoint, suggesting a partial duplication or more intricate structural event. OMKar determined that it is not possible to reconstruct both homologous chromosome pairs without either excluding or introducing additional calls. OMKar automatically inferred a single additional translocation edge between segments 48 and 66 and generated a complete karyotype incorporating all reported SV and CNV calls (Supplementary Fig. S6B-D).

### OMKar correctly reconstructs variations with partially missing calls

Ideally, unbalanced rearrangements are supported by both SV edges and by CNVs. However, the CNV call might be missed if the region is small, and SV call might be missed if the breakpoints lay in regions of low-complexity that are masked by the Bionano pipeline. OMKar reconstructs karyotypes with rearrangements that are supported only by SVs or only by CNVs. Specifically, it infers missing SVs by adding additional edges to make the breakpoint graph Eulerian (Methods). OMKar outputs inferred variants as lower confidence karyotype features.

We reanalyzed 40 deletions and 42 amplifications detected by OMKar that were cross-validated using complementary technologies. Of the deletions, 12 (30%) were reconstructed using only CNV calls, and 4 (10%) deletions were reconstructed using only SV calls. Among the amplifications, 15 (36%) amplifications were reconstructed using only CNV calls.

In prenatal sample 205, an inter-chromosomal duplicated insertion was previously reported using a combination of karyotyping and CMA: ins(14;2)(q32;q36.1q31.2) (105.159M; 221.205M-178.043M). OGM reported a high confidence inter-chromosomal SV call of Chr14: 105.159M to Chr2: 221.205M, with the correct orientation, but the other inter-chromosomal SV call was missing. It also reported a CN gain of the duplicated region of Chr 2. OMKar correctly inferred the missing SV call using support from the other SV and CNV calls, and was able to automatically reconstruct this rearrangement.

### OMKar identifies the genetic basis of previously diagnosed phenotype

The OGM samples were generated based on different usage modalities (Supplementary Table S2). Prenatal testing in 50 samples was performed, either because of an abnormality detected by non-invasive screening (44 samples) or because of elevated risk due to family history or advanced maternal age. In contrast, 28 of 34 unique postnatal samples presented with a clinical phenotype. The 63 parental samples contained individuals who had experienced miscarriage, had a higher probability of translocation, or suspicion of infertility.

Among the pre- and postnatal samples, 20 had a previously diagnosed Genotype-to-Phenotype (G2P) mechanism, largely obtained through a manual analysis of CMA and karyotyping. OMKar automatically identified all 20 G2P mechanisms through a correlation between reconstructed karyotypes and an intersection with the DDG2P database (Methods). These were all aneuploidies, with phenotypes including Triple-X, Jacobs, Turner Syndrome, and Down Syndrome. In contrast, for the (largely asymptomatic) parental samples, OMKar automatically and correctly reconstructed 9 of 10 translocations (missed one Robertsonian) to explain eight infertility cases and one clinically remarkable child. The last case (ID:1999) highlighted the importance of parental karyotyping using OMKar. The sample was unremarkable in cytogenetic karyoptying but carried a balanced translocation between the p-arms of Chr 4 and Chr 7, t(4;7)(3,903,798;6,881,853). This balanced translocation resulted in the inheritance of an unbalanced translocation. The child carried a deletion of Chr4: 0 - 3.9 Mbp, causal for the Wolf-Hirschhorn Syndrome (deletion of Chr4: 1.6 - 2.1 Mbp)(Wolf et al., 1965; Zollino et al., 2003).

Apart from the 10 cases, OMKar also identified a deletion, del(X) (31,614,556-31,831,572) in one parent (ID:19), which causes a monoallelic loss of the gene *DMD*, leading to Becker muscular dystrophy (BMD)(Ervasti et al., 1990). BMD can be inherited, and while the deletion was likely not causal for the observed infertility, it may have contributed in combination with other un-diagnosed factors.

All of these confirmatory diagnoses either involved large CNVs that could be resolved by CMA, or large translocation events that could be detected by karyotyping. We next investigated OMKar’s capacity to detect smaller SVs.

### OMKar reconstruction explains genetic basis of postnatal phenotypes

In addition to the larger variants discussed previously, after filtering (Section S3.1), OMKar reported 28 balanced (copy-neutral) SVs and 144 unbalanced SVs in 21 postnatal samples with missing G2P explanations. Importantly, OMKar also provided novel G2P explanations for 5 of 21 samples(Table 3).

**Table 3:** OMKar G2P explanations for undiagnosed postnatal cases. ID: Intellectual Disability, DD: Developmental Delay.

| Sample | Clinical Indication | Clinical Diagnosis | OMKar Genotype | OMKar G2P Prediction |
| --- | --- | --- | --- | --- |
| 2081 | Absent teeth, ID | ID | (new) *PAX8* deletion | Congenital Hypothyroidism non-goiterous type 2 |
| 2280 | Mild dysmorphic features, hypotonia, DD | Kabuki Syndrome; Unknown | (new) deletion in the middle of translocation, *WDFY3* + *HNRNPD* | Primary Microcephaly or macrocephaly with developmental delay; *HNRNPD*-related developmental disorder (monoallelic) |
| 2276 | Seizures, ID | Unknown | translocation interrupts *TANC2* | *TANC2*-related neurodevelopmental and psychiatric disorders |
| 2281 | DD | DD | transposition interrupts *MBD5* | *EHMT1*-like ID |
| 2282 | ID | ID | translocation interrupts *SOX5* | 12P12.5 Intragenic deletions associated with ID |

Two samples showed unbalanced events with a loss of genes important for neurodevelopment. For sample 2081, it was a deletion call (10.4 Mbp). The karyotype for sample 2280 was more complex, with a deletion (5.1 Mbp) in the middle of a translocated segment (segment 4c, Fig. 3A-C). Previously, only karyotyping had been performed for both cases, and neither deletion was detected.

**Figure 3.**
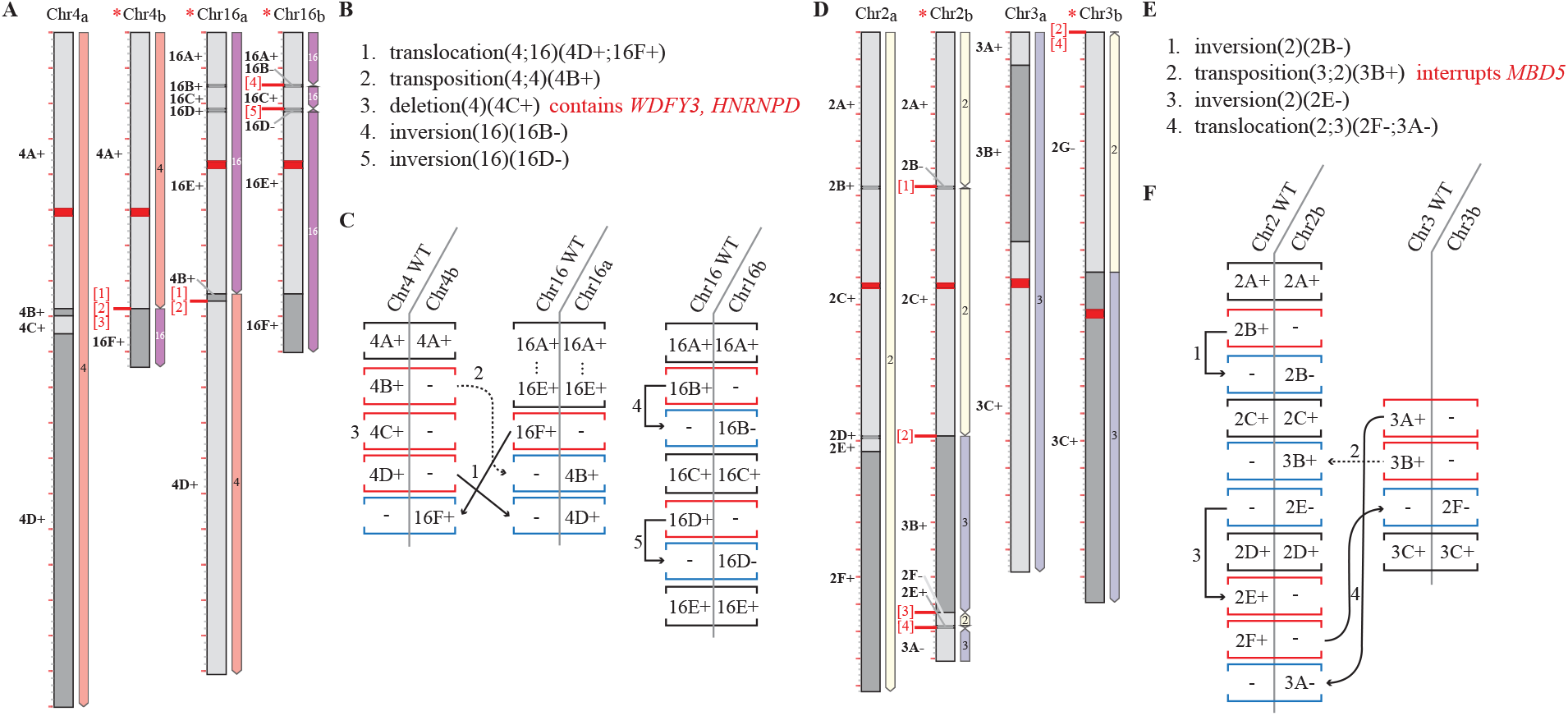
OMKar reconstructions in two postnatal samples: 2280 (panels A-C) and 2281 (panels D-F). The karyograms (panels A, D) and the ISCN-formatted description (panels B, E; slightly altered for exposition) were automatically generated by OMKar. The karyograms displayed show the ‘segment view’ for easier referencing. Panels C, F describe the SV interpretation process after path decomposition, with black brackets indicating concordant blocks, red indicating deletion blocks, and blue indicating insertion blocks.

Among 13 balanced events reconstructed by OMKar in these 5 samples, boundaries in 3 samples (2276, 2281, and 2282, see Table 3) interrupted a neurodevelopmental gene. Because precise coordinates of translocation were not reported via karyotyping or CMA, a genotypic basis was not previously diagnosed. Sample 2281 in particular, illustrates the power of OMKar’s reconstruction of a complex karyotype (Fig. 3D-F). The reconstruction revealed a transposition of a Chr 3 segment on to Chr 2, interupting the *MBD5* gene between segments 2C and 2D. The karyotype additionally included two inversions and a translocation resulting in a highly rearranged chromosomal cluster with no change in copy number.

## Discussion

Karyotyping remains an essential tool in the diagnosis of constitutional genetic disorders, particularly those arising from large chromosomal rearrangements such as aneuploidies, translocations, and complex structural variants. While traditional methods such as cytogenetic karyotyping, FISH, and microarray have long served as the standard for detecting these abnormalities, they are constrained by limited resolution, manual labor intensity, and an inability to detect balanced rearrangements and novel variations with high precision. In contrast, genomic technologies (whole exome/genome sequencing) are very precise, but do not easily provide chromosome level characterizations. Long-read technologies are currently too expensive for clinical use. Optical Genome Mapping (OGM) thus represents a happy medium, offering a medium-resolution, robust alternative capable of detecting a broader range of structural variations (SVs) in clinical settings(Smith et al., 2022; Valkama et al., 2023). Recent clinical studies have shown OGM have high concordance (99.5%) with standard-of-care (SOC) methods over 1,000 samples, with increased detection rate of pathogenic or likely-pathogenic variants(Iqbal et al., 2023; Broeckel et al., 2024). OGM is also recently incorporated into the International System for Human Cytogenomic Nomenclature (ISCN)(Hastings et al., 2024). For these reasons, we developed our tool starting with OGM data. Our method, OMKar, bridges the gap between low-resolution techniques like cytogenetics and high-resolution sequencing methods by capturing large-scale rearrangements, but also combining the information into an automated karyotype inference. It identifies key structural abnormalities such as balanced translocations, inversions, and duplications. The ability to automate karyotyping through OMKar not only reduces the manual workload but also enhances the speed and scalability of the analysis. This enables clinicians to analyze large datasets, improving diagnostic accuracy and potentially leading to faster treatment decisions.

In developing OMKar, we faced a significant challenge of resolving conflict between SV and CNV calls. Such conflicts were caused by either having one of the calls with high confidence while the other call was missed or masked or when SV and CNV call boundaries were not identical. OMKar is designed to infer variations with partially missing or conflicting calls and resolve boundaries. Future developments in OGM technology and in algorithmic reconstruction should reduce these conflicts, resulting in higher confidence calls.

OGM has a practical resolution limit for CNV detection, with high-confidence calls generally restricted to events ≥ 100–150 kb. In OMKar, we apply a conservative filtering threshold of 200 kb to exclude low-confidence CNV calls, thereby enhancing the specificity of the breakpoint graph and reducing computational complexity during reconstruction. Notably, smaller CNVs that overlap with SV calls are reintegrated into the analysis, and SVs with only partial CNV support are flagged as lower-confidence features. This approach preserves biologically relevant variation while maintaining the accuracy and tractability of the method, and reflects the current resolution limitations inherent to OGM platforms.

OGM allows genome-wide detection of large SVs with high sensitivity, including balanced events like translocations and inversions that are often missed by sequencing. However, OGM cannot detect single nucleotide variants (SNVs) or small indels, and it has reduced sensitivity for small CNVs (*<*100 kb), centromeric regions, and mosaicism. Despite these limitations, OGM fills a critical gap between cytogenetics and sequencing in clinical genomics.

OMKar utilizes the Bionano pipeline for SV calling, which in turn is based on searching the human reference genome. With the availability of multiple genomes, SV calling can be improved using assembly graphs or even de Bruijn graphs as the reference(Mukherjee et al., 2019, 2021; Alipanahi et al., 2016; Leinonen and Salmela, 2020; Lin and Pop, 2011). Future work will seek to incorporate these technologies to improve SV calling and karyotyping.

OMKar (and OGM), while highly effective for most structural variants, shows reduced sensitivity in detecting mosaic chromosomal abnormalities, events occurring in regions of low complexity and segmental duplications that can lead to non-allelic recombination such as Robertsonian translocations. Mosaicism, characterized by variation in chromosomal numbers within different cell populations, also pose a challenge for OGM, which is primarily focused on large, stable genomic rearrangements. Further refinement of OGM technology and the tools will be needed to broaden its applicability in clinical settings.

OMKar showed a performance gap between terminal and non-terminal structural variation. Previous results from FISH screening suggest that a small number (5-10%) of developmental disorders that lead to intellectual disability are due to “cryptic telomeric rearrangements”(Moeschler and Shevell, 2006). However, this may be an underestimate because subtelomeric rearrangements are often seen as *de novo* variants(Luo et al., 2011). Our future research will focus on algorithms for identifying telomeric abnormalities, including ring chromosomes(Mostovoy et al., 2024).

OGM technologies cannot currently detect variations within centromeres or the short arms of acrocentric chromosomes(BionanoGenomics, 2018, 2024). In particular, Robertsonian translocations involve rearrangements of the short-arms of acrocentric chromosomes are not currently detected by OGM. Recent studies have suggested the use of long-read technologies like Oxford Nanopore for detecting them(Mostovoy et al., 2024). Because the core of OMKar algorithm, which includes the building of Eulerian graphs followed by path extraction, is agnostic of a specific sequencing technology, OMKar should be easily adapted to other technologies. As more datasets describing these events are made available on different sequencing platforms, we plan to develop karyotyping tools for those platforms.

In conclusion, OMKar has demonstrated significant potential in automating and improving the accuracy of karyotyping using OGM data. It offers a robust, scalable, and high-resolution approach to detecting constitutional genetic abnormalities, though ongoing improvements are necessary to fully address its limitations. As the tool continues to evolve, it could become an increasingly important method for research and clinical diagnostics, complementing and potentially surpassing traditional methods in terms of accuracy and efficiency.

## Methods

### Data pre-processing

OMKar filters the SV and CNV calls to ensure data quality and relevance of the SV to karyotyping. OMKar utilizes the default Bionano pipeline thresholds for calling CNVs and SVs to ensure that only reliable variants are used in karyotype reconstruction. The Bionano pipeline maintains a database of regions that are observed with structural or copy number variation, and masks genomic regions that are frequently seen in normal samples. OMKar filters out CNV calls in the masked regions if they do not have supporting SVs. Finally, OMKar filters CNVs smaller than 200 kbp, and those are reincorporated as local changes after karyotype reconstruction. (see Supplementary Section S3.1).

Breakpoints from SV calls are further processed, sorting them by chromosomal and genomic coordinates and merging adjacent breakpoints within a 50 kbp window, while also ensuring precise representation of structural variations by splitting CNV segments when breakpoints occur within their boundaries. The result of this pre-processing is a partitioning of each chromosome into a minimal number of segments, so that (a) each segment has a nearly uniform copy-number, (b) all breakpoints link only the end-coordinates of segments, (c) segments span all regions of reference chromosomes with copy number ≥ 1.

### Breakpoint graph construction

We use the genome segment partitioning to generate a breakpoint graph(Alekseyev and Pevzner, 2009) *G*(*V, E*_*s*_ ∪ *E*_*r*_ ∪ *E*_*b*_). Each vertex *v* ∈ *V* corresponds to the end-coordinate of a segment. The set of *segment-edges* is defined using *E*_*s*_ = {(*u, v*) s.t. *u* and *v* are canonically the head and tail-node of the same segment}. The *non-segment* edges include two sets. First, the set of *reference-edges, E*_*r*_ = {(*v, u*)} where the head-node *u* of a segment is *adjacent in reference coordinates* to the tail-node *v* of another segment, and second, *breakpoint-edges* (*u, v*) ∈ *E*_*b*_ joining vertex *u* to a *non-adjacent* vertex *v*. By definition, each vertex *v* is incident on exactly one segment-edge, at most one reference-edge, and possibly multiple breakpoint edges denoted by *E*_*b*_[*v*].

### Smoothing Edge Multiplicities to generate an Eulerian graph

We use an Integer Linear Programming (ILP)(Schrijver, 1998) formulation to constrain the copy number of each genomic segment. Let *c*_*v*_ denote the copy number assigned to the segment-edge incident on vertex *v*. The ILP assigns copy numbers *r*_*v*_ ≥ 0 to the reference edge, *s*_*e*_ ≥ 0 to each edge in *E*_*b*_[*v*] and auxiliary values *x*_*v*_ while enforcing the following constraints:

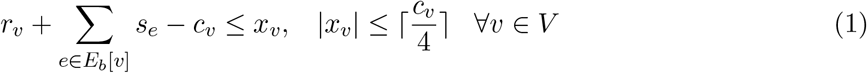

These two constraints ensure that the sum of copy numbers of outgoing edges from a segment is not greater than the segment’s assigned copy number. Moreover, if a reference-edge incident on *v* is supported by a contig alignment that spans the adjacent segments, then *r*_*v*_ ≥ 1. In order to make the graph Eulerian, we need to reduce the number of vertices with odd degrees. For this purpose binary parameter *o*_*v*_ is defined as follows:

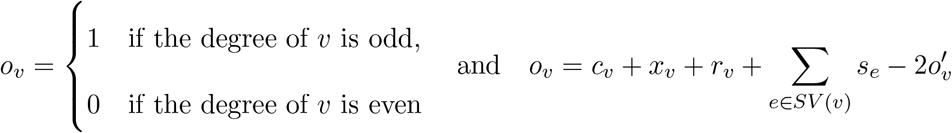

In which 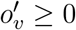 is an auxiliary variable added to the ILP. We minimize an objective function

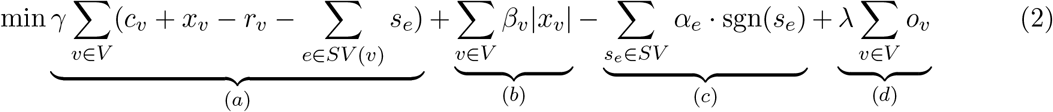

where (a) penalizes for the discrepancy between observed plus slack copy number of segment edges and the total copy number of adjacent reference and breakpoint edges; (b) penalizes for using non-zero slack; (c) provides a reward for using breakpoint edges at least once and (d) penalizes for having odd degree vertices. The actual implementation linearizes the non-linear terms (Supplementary Section S3.2). The objective includes the parameters *γ, α*_*e*_, *β*_*v*_, *λ* which were set empirically(Supplementary Section S3.3, Supplementary Table S4).

OMKar uses a fixed set of parameters designed to detect common structural variants, without dataset-specific tuning. These parameters were applied consistently across 38 simulated genomes and 154 clinically diverse samples from ten independent sites, demonstrating robustness and generalizability despite variability in sample type and data quality.

### Computing Eulerian tours

Following the estimation of edge multiplicities, we utilize a Breadth-First Search (BFS) algorithm to identify all connected components within the graph, each component representing a chromosome cluster. For each connected component, denoted as *C*, our approach initiates with the identification of vertices that represent the telomeric regions of chromosomes. If *C* contains odd-degree non-telomeric vertices, we connect such pairs using dummy edges to transform *C* into an Eulerian structure. Algorithm 1 (Supplementary Section S3.4) is used to compute Eulerian tours originating from one of the telomeric vertices.

### Chromosomal Segregation and identification

In this formulation, each chromosome is a subpath with alternating segment and non-segment edges. A connected component may incorporate multiple chromosomes. Therefore, in Algorithm 1, we compute Eulerian paths that force an alternation between segment and non-segment edges. In case the only possible transition from segment edge (*u, v*) is to the segment edge (*v, u*), this represents a boundary between homologous chromosome pairs, and the subpaths are split at node *v*.

Eulerian decomposition is not unique, and multiple decompositions may exist. Let us assume that after path segregation, we have a set of sub-paths *P* = *P*_1_, *P*_2_, Now, consider two paths *P*_*i*_ and *P*_*j*_ that share the same segment *s*. In that case, a ‘crossover transition’

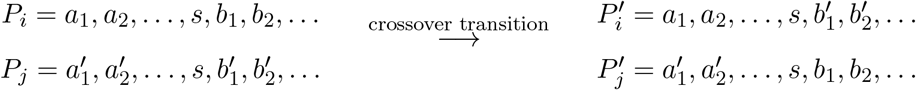

would generate another valid Eulerian path decomposition. We use this idea to heuristically refine the chromosomes based on known biology. Specifically, OMKar counts the number of centromeres in each path. If there exists a pair of paths–one containing two centromeres with at least one segment *s* in between and a second chromosome containing segment *s* but zero centromeres–OMKar performs a crossover transition on *s* to ensure that both paths now contain a single centromere. Finally, to standardize orientation, we flip the chromosomes so that they are all oriented in the p-to-q direction. Each chromosome’s orientation is identified by the centromeric segment, or, if it is acentric, by the majority orientation of all segments.

### Event Interpretation

Structural variants have somewhat conflicting definitions within the Genomics and Cytogenetics community. We developed an Event Interpretation module to describe SVs using the International System for human Cytogenomics Nomenclature (ISCN), described in Supplementary Table S5. OMKar automates the interpretation as follows (See also, Supplementary Section S3.5): It aligns structural variations (SVs) in reconstructed chromosomes with their wild-type (WT) counterparts, identified by centromeric or segmental makeup. It uses the Longest Common Subsequence algorithm to create blocks and classify them as concordant, insertion, or deletion. Adjacent blocks with the same classification and contiguity are combined. Each insertion or deletion block is assigned an ISCN based on unique block-type signatures, where indel size allowance determined via simulation (Supplementary Table S5, Supplementary Fig. S7). The system favors interpretations involving single, complex SVs over multiple simpler ones, providing a comprehensive explanation for the observed chromosomal deviations (Supplementary Section S3.5).

### Report

Based on the interpreted SVs, disrupted genes that are present in the DDG2P database(Thormann et al., 2019) are reported. For balanced SVs, we looked at the boundaries within a resolution of 5 kbp for disrupted genes that might lead to a loss of function (resolution determined empirically with simulations, Supplementary Section S4). For unbalanced SVs, we looked at the entire affected regions, for either gain or loss of gene product. Lastly, the allelic (monoallelic/biallelic) and mutational (loss/gain/altered gene product) requirements are used to filter for disrupted gene output and phenotype prediction.

An HTML report is compiled for ease of reading. It includes the decomposed paths (chromosomes), the corresponding visualization of the chromosome in both cytoband and segment views (Supplementary Section S3.6), the interpreted SVs under the ISCN language, and the disrupted developmental genes.

### KarSim module for simulating karyotypes

The KarSim module (Supplementary Section S5.1, Supplementary Fig. S8) generates a molecular karyotype file, a FASTA file, and a history log of events for downstream use, including KarCheck comparisons, while also allowing for the simulation of different sequencing technologies.

#### Usage in Simulation tests

Random karyotypes were generated to simulate a common genetic disorder from Decipher Database’s CNV syndromes(Firth et al., 2009), followed by 7 to 14 random de-novo SVs from Supplementary Table S6. Certain genomic regions, including centromeres and telomeres, were masked during the analysis to ensure accurate structural variant placement. More details on the masked regions can be found in the Supplementary Section S5.1. SVs were placed with breakpoints at least 50 kbp from masked regions, ensuring no segments smaller than 50 kbp were generated.

After generating the parameterized-random molecular karyotypes, simulated data was processed with OMSim(Miclotte et al., 2017) to generate OGM molecules with added noise. The standard Bionano Solve pipeline (v3.7)(BionanoGenomics, 2018) was applied to compute CNVs, SVs, and contig alignments, which were then used as input for OMKar to reconstruct the final virtual karyotype. Full details on the simulation process and parameters can be found in the Supplementary Section S5.3.

### The KarCheck module for comparing karyotypes

Karyotypes from the simulation (*K*_*t*_) are compared with reconstructed karyotypes (*K*_*r*_) using the KarCheck module (Supplementary Section S6, Supplementary Fig. S8). Preprocessing is initially applied to *K*_*t*_ and *K*_*r*_ to divide chromosome groups into clusters. To achieve comparability, segments are further divided so both karyotypes share the same set of segments (Supplementary Section S6.1). For each chromosome cluster, three metrics are reported: 1) chromosome count concordance, 2) Jaccard similarity of SV edges, and 3) Jaccard similarity of CN.

#### SV similarity computation

SV similarity computation is performed by comparing non-segment edges in the pair of chromosome clusters. The edges are matched (allowing for some tolerance, analyzed in Supplementary Section S4), and a Jaccard similarity score (Intersection over Union) is computed to measure similarity (Supplementary Section S6.2). OGM reads do not map with high confidence in the telomere, acrocentric p-arm, and acrocentric centromere regions. Therefore, these prefix/suffix regions were excluded in simulations and during SV similarity computations.

#### Copy Number similarity comparison and metrics

CN similarity comparison is done by binning the whole genome (excluding prefix/suffix masked region) into spanning, non-overlapping bins of 50 kbp, with a tolerance of +/-100 bp (exact size chosen to maximize the size of the last bin on the chromosome). Each bin is used to store the average CN within that region, and a bin is termed “with CNV” if deviates more than 0.05 from diploid (chosen based on OGM’s resolution). A Jaccard similarity is computed between the bins with CNV. (Supplementary Section S6.3).

## Data Availability

The optical genome mapping (OGM) datasets analyzed in this study are publicly available from the European Nucleotide Archive (ENA) under accession number (PRJEB90248), and the European Genome Archive (EGAS00001008245). All raw data used for karyotype reconstruction are included in this submission. Metadata and analysis files are organized per sample, and detailed instructions for accessing and interpreting the data are provided in the repository.

https://www.ebi.ac.uk/ena/browser/view/PRJEB90248

## Ethics Review Statement

This study involved human participants and was approved by the Ethics Committees of the Medical University of Vienna (ethical code: 2229/2019) and Keçiören Teaching and Research Hospital (ethical code: 2012-KAEK-15/2083). The study adhered to the principles outlined in the Declaration of Helsinki. Informed consent was obtained from all participants prior to their inclusion.

Additionally, the study was conducted in accordance with the Declaration of Helsinki and received approval from the Institutional Review Boards of Western IRB – Copernicus Group (WCG) under study numbers 20203726 and 20212956. This approval included provisions for informed consent or waived authorization for the use of de-identified, banked samples for research purposes. All protected health information (PHI) was removed, and data were anonymized (coded and double-blinded) before accessioning for the study.

## Software Availability

The OMKar source code is publicly available under an open-source license at: https://github.com/siavashre/OMKar. The repository includes detailed documentation, example datasets, and scripts for reproducing key results presented in this study. To facilitate reproducibility and ease of deployment, we also provide a Docker image of OMKar that can be executed on Linux, macOS, Windows, or cloud environments without manual dependency installation. A Conda version is also provided.

## Data Access

The optical genome mapping (OGM) datasets analyzed in this study are publicly available from the European Nucleotide Archive (ENA) under accession number PRJEB90248, and the European Genome Archive (EGAS00001008245). All raw data used for karyotype reconstruction are included in this submission. Metadata and analysis files are organized per sample, and detailed instructions for accessing and interpreting the data are provided in the repository.

## Acknowledgments

This work was supported by the National Institutes of Health (NIH) grant R01GM114362. VB is a co-founder and scientific advisory board member of Boundless Bio, Inc. (BBI) and Abterra Inc., holding equity in both companies. BBI and Abterra were not involved in this research.

We thank Gautam Kathir for developing the initial HTML report and Christopher Day for discussions on database-related aspects. We also acknowledge the authors of the two multicenter studies (PMID: 36828597, 38211722, 39032820) for providing 98 clinical OGM datasets used in the evaluation of OMKar.

Some de-identified data used in this study originated from a study sponsored by Bionano Genomics, specifically from the “Validation of Optical Genome Mapping for the Identification of Constitutional Genomic Variants in a Postnatal Cohort” study (NCT05295277; https://clinicaltrials.gov/study/NCT05295277?term=Optical%20Genome%20Mapping&rank=6).

S.R.D. and Z.J. designed and implemented all software and analyses for this study, contributed to result interpretation, and co-wrote the manuscript. J.E. assisted with software implementation. J.H. and N.M. provided data and contributed to the interpretation of cytogenetic results. N.G.L. and J.N. provided clinical data. A.H. and A.C. provided data and contributed to the interpretation of cytogenetic results. A.W.C.P. provided data, contributed to cytogenetic interpretation, and codesigned the study scope. P.D. provided clinical data, contributed to result interpretation, and assisted in manuscript writing. V.B. co-designed the study scope, help design the algorithms, contributed to result interpretation, and co-wrote the manuscript.

## Supplementary Figure Captions

**Supplementary Figure S1:**
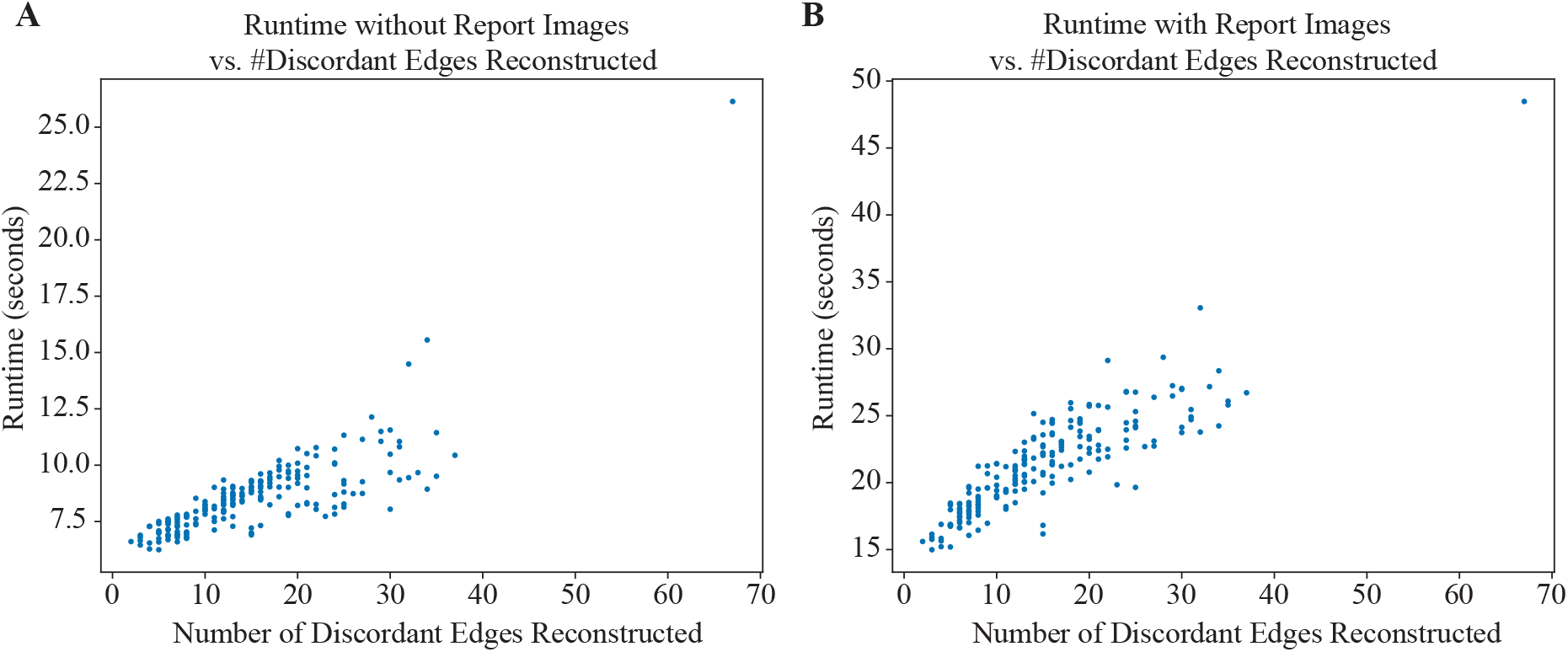
Runtime analysis. Runtime of OMKar was collected on both simulation and clinical samples A) without rendering the visualization images in HTML report, and B) with rendering the report images.

**Supplementary Figure S2:**
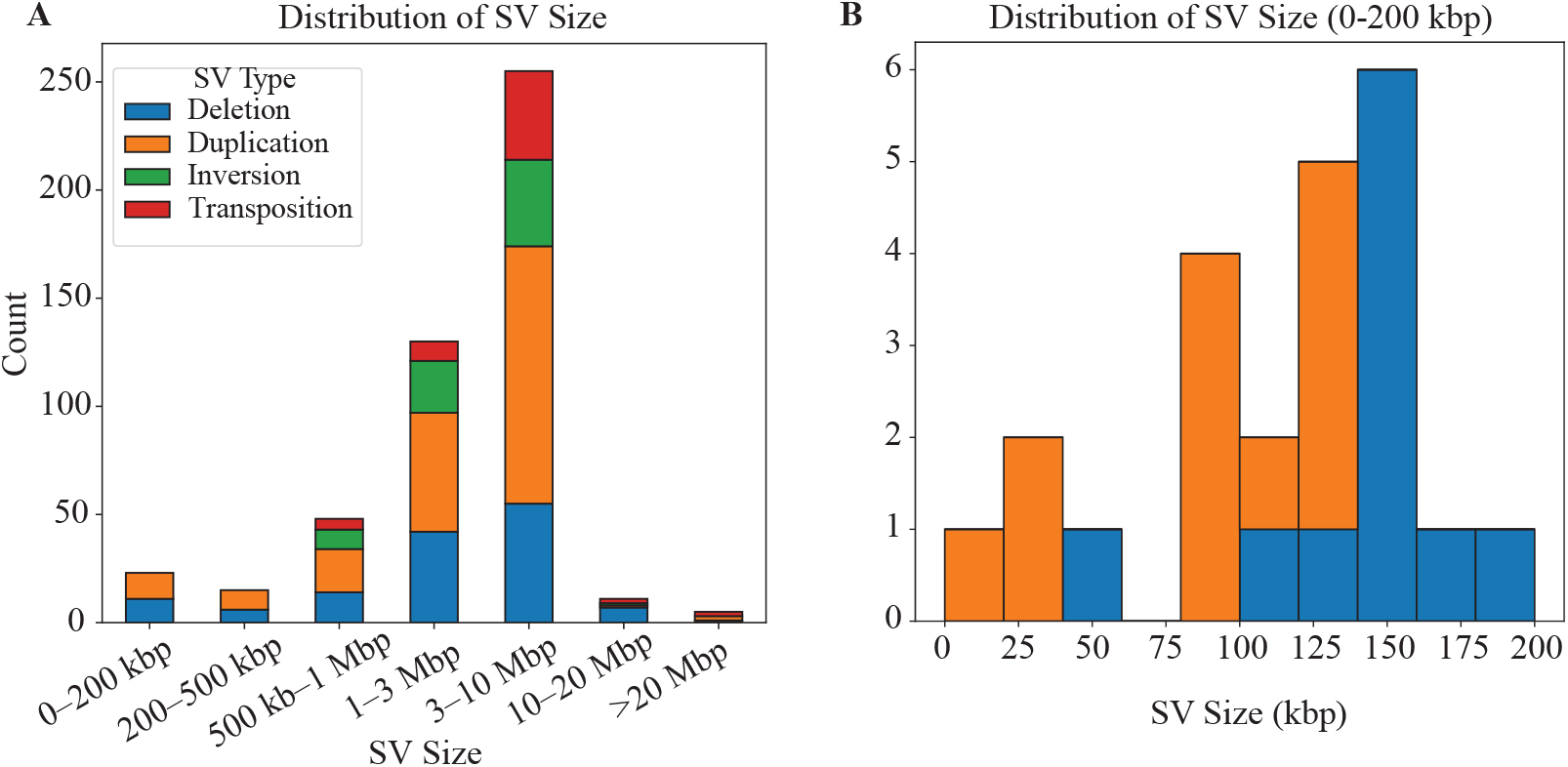
SV size distribution. A) Binned SV size distribution of all samples evalulated. B) Detailed size distribution between 0 and 200 kbp.

**Supplementary Figure S3:**
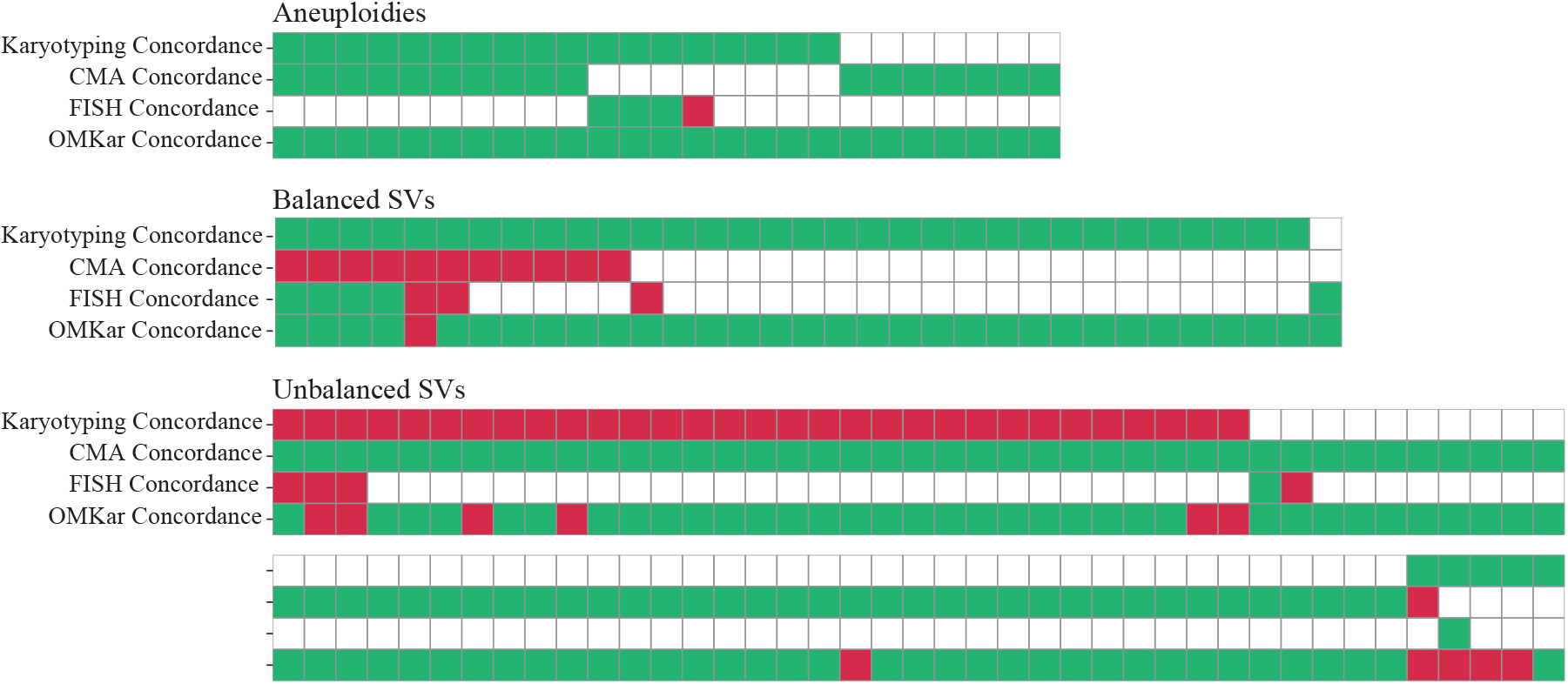
Validation of OMKar reconstruction against clinical annotation by cytogeneticists using karyotyping, CMA, and/or FISH. OGM and OMKar was applied to all samples, but not all technology was applied on all samples. Each column represents a clinical sample. The color code in the cells explain the concordance of the applied technology. White: the technology was not applied; Green: concordance; Red: Missed. Variations were grouped into A) aneuploidies, B) balanced structural variations, and C) unbalanced structural variations. Note, the unbalanced structural variations take up two sets of rows.

**Supplementary Figure S4:**
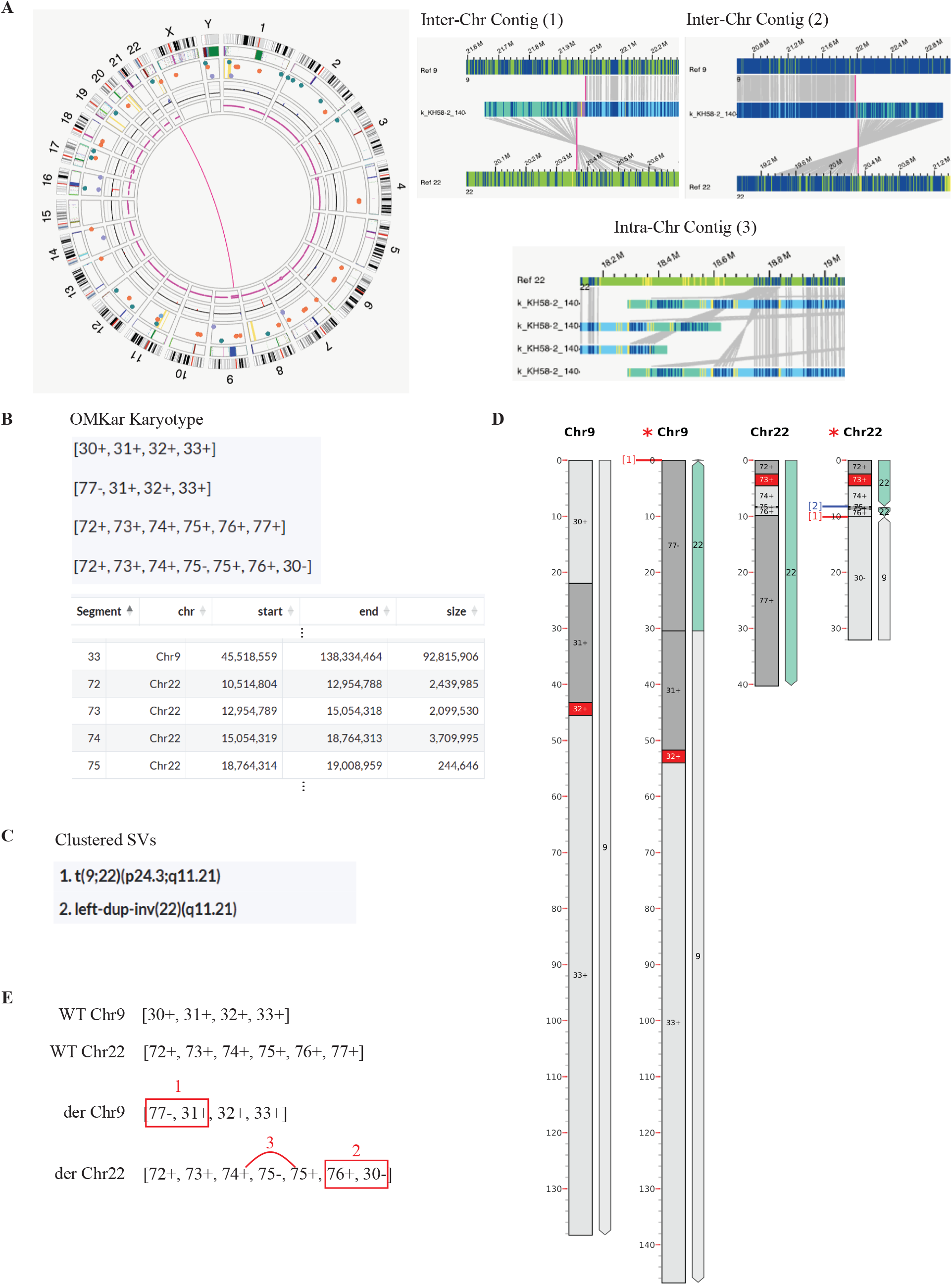
Comparison of Bionano Access and OMKar Outputs (Postnatal 1404). Postnatal sample 1404 contains a balanced reciprocal translocation between Chromosomes 9 and 22, as well as a 240 kbp left-duplication inversion on Chromosome 22, located approximately 1.3 Mbp upstream of the translocation breakpoint. (A) The inter-chromosomal translocation is called and shown on the Circos plot from Bionano Access. The two translocation contigs of the same orientation are shown on the right, alongside the contigs supporting the duplication inversion call. (B) The OMKar output on the same data gives a clear description of the karyotype as a sequence of chromosomal segments (e.g., …74+ 75-75+ 76+ 30-, in derivative Chromosome 22), where each segment number corresponds to a distinct genomic interval. (C) A chromosomal annotation of the OMKar reconstruction shows the canonical balanced translocation event, forming the derivative Chromosomes 9 and 22. In addition, there is a left duplication inversion on derivative Chromosome 22 upstream of the translocation. (D) As part of the automated OMKar report, the clustered SVs are also shown in ISCN format. (E) The automated chromosomal ideogram corresponding to the reconstruction is included on the right, with the chromosomal segments and the location of the clustered SV labeled.

**Supplementary Figure S5:**
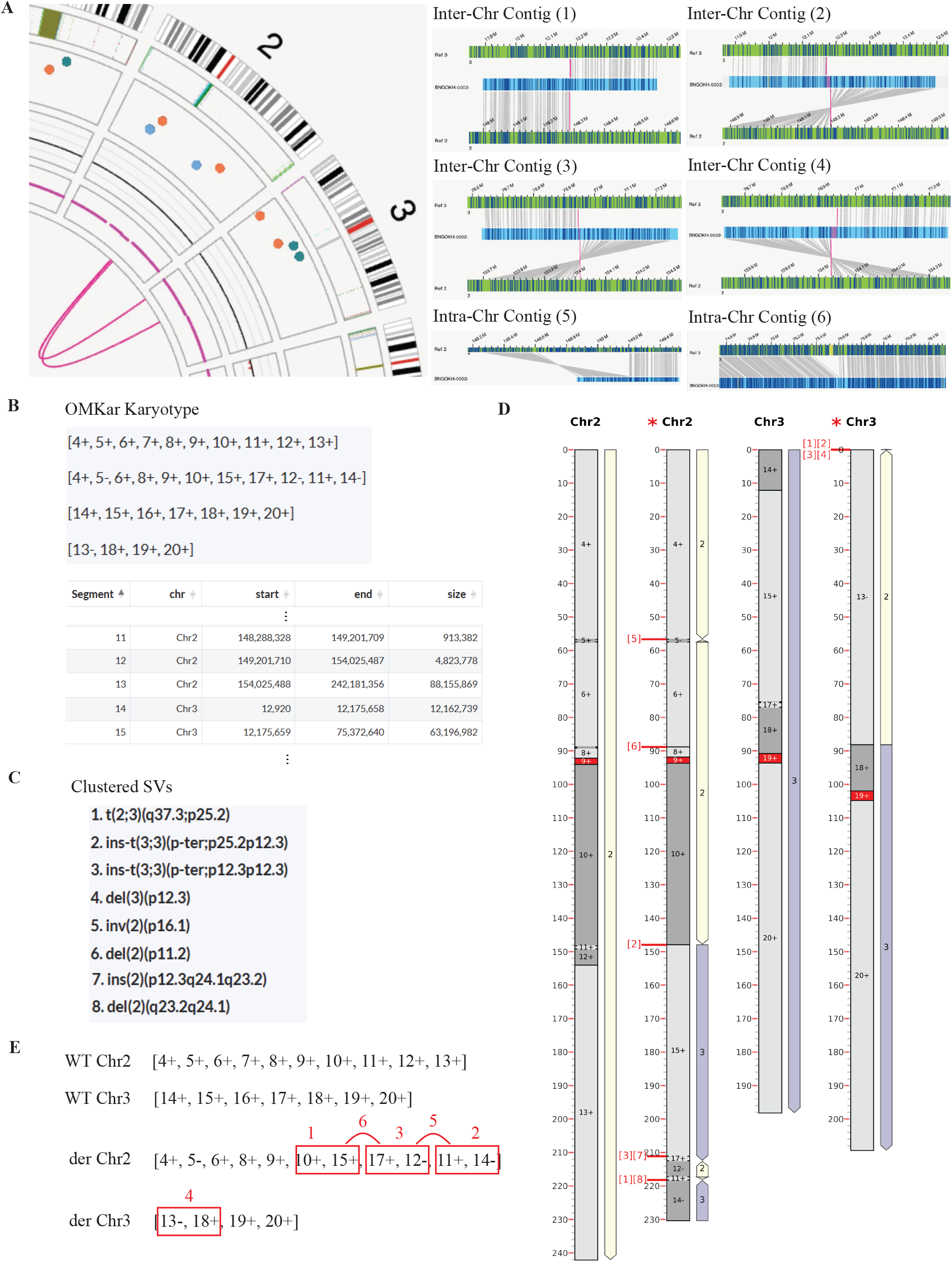
Comparison of Bionano Access and OMKar Outputs (Postnatal 2281). Postnatal sample 2281 contains a set of complex translocations called between Chromosomes 2 and 3. (A) There are four translocations called but shown only as two distinct curves in the Circos plot from Bionano Access because of overlaps. The detailed contig alignments from the Genome Browser View are shown on the right. The four translocations occur at two distinct breakpoint regions, with each region involving a pair of translocations in different orientations. In addition, an inversion and a deletion call were also found in the region. (B) The OMKar output on the same data gives a clear description of the karyotype as a sequence of chromosomal segments (e.g., …10+ 15+ 17+ 12-… in derivative Chromosome 2), where each segment number corresponds to a distinct genomic interval. (C) A chromosomal annotation of the OMKar reconstruction shows the complexity of this karyotype which is a non-canonical translocation event that explains all 4 translocations reported by Bionano. The derivative Chromosome 2 starts with an inversion of segment 5 and a deletion of segment 7, followed by the translocation to segment 15, a deletion of segment 16, translocation back to the reversed segment 12, an inversion to segment 11, and translocation to the inverted segment 14. The derivative Chromosome 3 is a canonical translocation of the inverted segment 13 to segment 18. (D) As part of the automated OMKar report, the clustered SVs are also shown in ISCN format. (E) The automated chromosomal ideogram corresponding to the reconstruction is included on the right, with the chromosomal segments and the location of the clustered SV labeled.

**Supplementary Figure S6:**
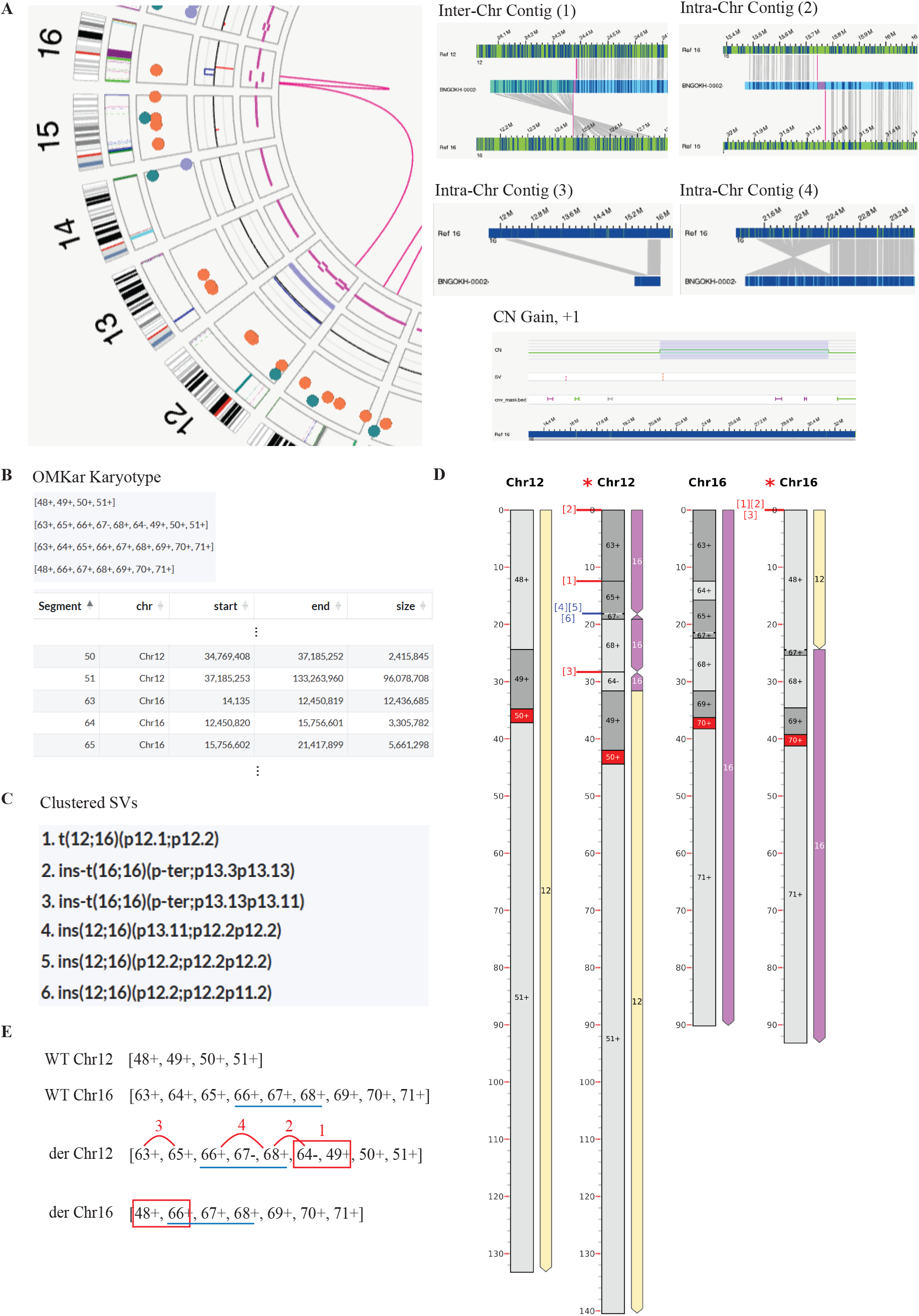
Comparison of Bionano Access and OMKar Outputs (Postnatal 2282). Postnatal sample 2282 contains a set of complex translocations called between Chromosomes 12 and 16. (A) One inter-chromosomal translocation, one intra-chromosomal translocation, one deletion, one inversion, and one copy number gain were called and shown in the Circos plot from Bionano Access. The detailed contig alignments and CNV region from the Genome Browser View are shown on the right. (B) The OMKar output on the same data gives a clear description of the karyotype as a sequence of chromosomal segments. (C) A chromosomal annotation of the OMKar reconstruction shows the complexity of this karyotype which is a non-canonical translocation event that explains all SV calls from Bionano and the CNV call that was not entirely aligned with the SV calls. The derivative Chromosome 12 starts on the p-arm of Chromosome 16 with a deletion of segment 64, followed by an inversion of segment 67, the intra-chromosomal translocation to the inverted segment 64, and the inter-chromosomal translocation to Chromosome 12. The derivative Chromosome 16 starts on the p-arm of Chromosome 12 and it contains the translocation to Chromosome 16 with the amplified segments 66, 67, and 68. (D) As part of the automated OMKar report, the clustered SVs are also shown in ISCN format. (E) The automated chromosomal ideogram corresponding to the reconstruction is included on the right, with the chromosomal segments and the location of the clustered SV labeled.

**Supplementary Figure S7:**
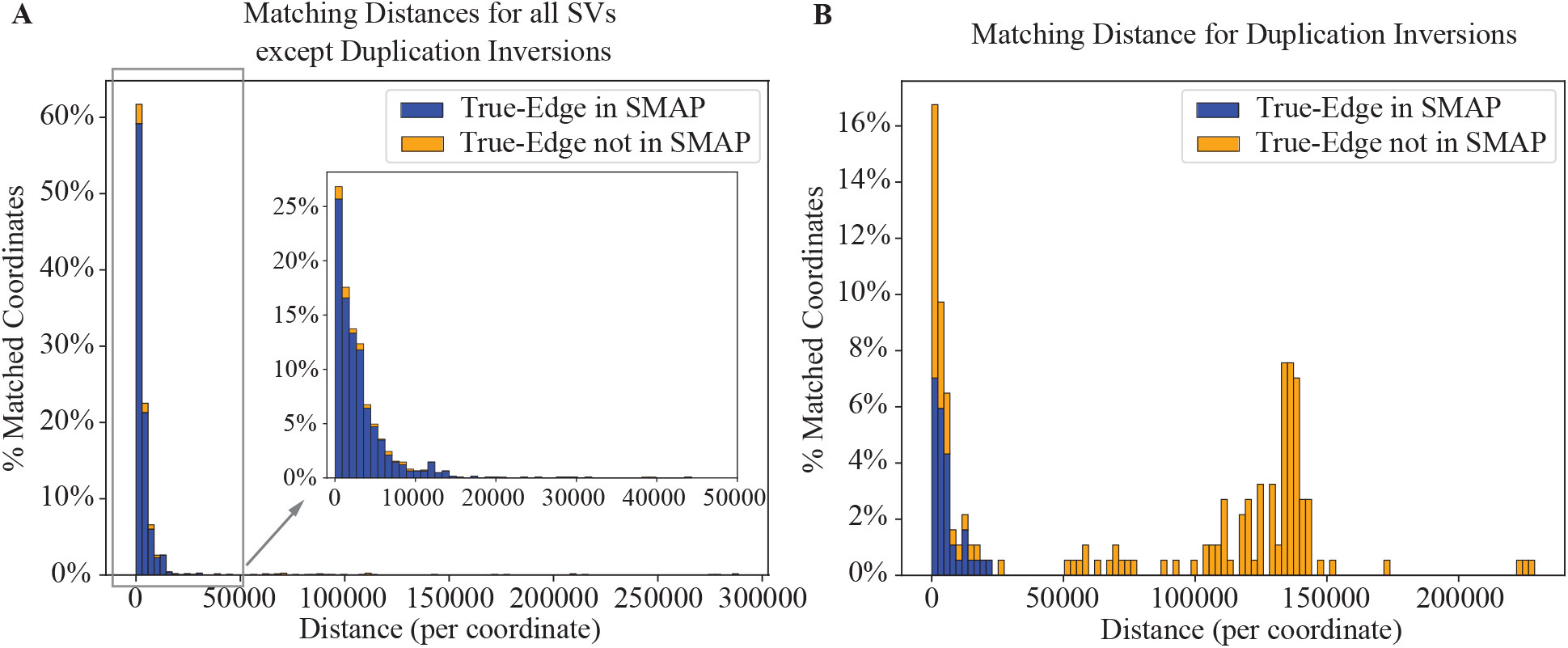
Matching distances per breakpoint coordinate. The breakpoint accuracy of OMKar is analyzed by comparing OMKar reconstructions against the simulated truth karyotypes. The KarCheck SV-edge matching breakpoint distance allowance was relaxed to 300 kbp to generate these plots. For each SV, the distance for each of the two breakpoint coordinates is computed separately. For better exposition, the SVs were partitioned into A) all SVs except Duplication inversions and B) duplication inversions.

**Supplementary Figure S8:**
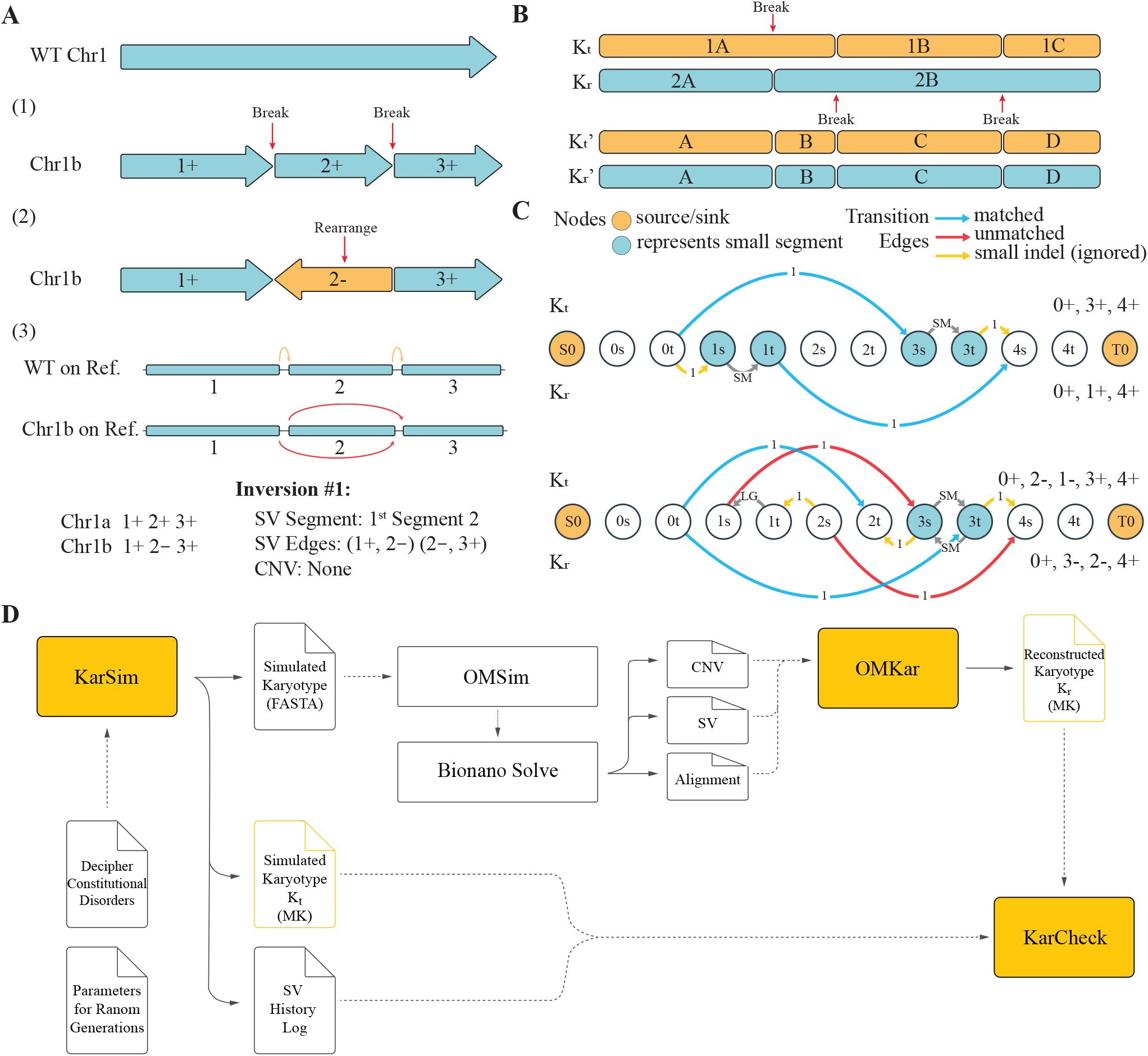
KarSim and KarCheck modules. A) Implementation of the KarSim for simulating karyotypes; B) Preprocessing module of KarCheck creates matching breakpoints between two karyotypes, allowing for direct comparisons. C) KarCheck Matching. Segments are annotated by size as being SM (small) or LG (large), and D) the complete simulation and validation pipeline for OMKar.

## Supplementary Table Captions

Supplementary Table S1: **Simulated terminal SV edge recall by event types.** Recall of SVs with breakpoint in the peri-terminal region. Different matching allowance was used in the KarCheck module to observe the reconstruction breakpoint distance from the truth SV breakpoint.

Supplementary Table S2: **Clinical sample information.** Includes information on each clinical sample. Sample ID: consistent ID as addressed in the paper, and it also distinguishes sample cohort; Test Site: location the sample was sequenced; Replicate Info: biological replicates of the sample, if applicable; Clinical Phenotype/Risk: clinical documentation of the reason this individual was tested; SOC G2P Prediction: clinical G2P diagnosis based on SOC methods (excluding OGM and OMKar); OMKar G2P Prediction: G2P diagnosis based on OMKar reconstruction; “Method” Performed: wether an SOC testing procedure was performed on this individual.

Supplementary Table S3: **Clinical sample variant information.** Includes information on each structural variation as identified by different techniques. The information includes the following. Cluster ID of the chromosome cluster for the SV; Cluster Chroms lists the chromosome groups within this chromosome cluster; Structural Variation: clinical annotation of this SV, written in ISCN annotation, based on SOC data only. Columns E-H describe the results using karyotyping, CMA, FISH, and OMKar. Columns I-L describe the concordance of the calls against the annotation. Column N describes reason why OMKar missed this particular variation, if applicable. Column O lists the G2P diagnosis based on OMKar reconstruction.

Supplementary Table S4: **OMKar parameters and tuning guidelines.** Summary of key parameters used in OMKar, including their default values, roles, and whether tuning is recommended for different datasets.

Supplementary Table S5: **Block signature for each structural variation.** Variations are listed in the order of finding. INS: insertion, DEL: deletion, CONC: concordant.

Supplementary Table S6: **Simulated structural variations.** (Canonical Structural Variants), where the two WT chromosomes are [1+ 2+ 3+] and [4+ 5+ 6+].

## Supplementary Material

### S1 OMKar Report

For ease of use, OMKar optionally compiles a report that includes 1) the observed chromosomal segment lists with reconstructed karyotype, 2) a list of interpreted events using ISCN notation, 3) a visualization of each corresponding chromosome with cytoband and event labels, and 4) a table of important genes that are near the breakpoints of an event or have copy number (CN) alteration. An html version of the report is prepared for easy viewing.

By default, OMKar outputs a molecular karyotype in a text format to unambiguously describe the karyotype as follows: It first lists all defined segments across the reference genome. For each segment, the following information is provided: segment number, chromosome, start and end coordinates, and the graph nodes representing the segment. Each segment is represented by two nodes, connected by a segment edge. All segments are forward-oriented (i.e., the end coordinate is greater than or equal to the start coordinate) and are sorted by chromosome groups (from 1 to Y) and increasing coordinates. The segments are non-overlapping, with no gaps between them, although telomeric regions of the reference genome may be excluded.

Following the segment definitions, OMKar reports the reconstructed paths that represent a karyotype. Each path consists of a list of segments in the format “Path number = segment number followed by direction.” The segments are traversed either in the forward (‘+’) or reverse (‘-’) direction, where ‘+’ indicates traversal from the start to the end of the segment, and ‘-’ indicates traversal from the end to the start of the segment. For example, “Path1 = 1+ 2+ 3-” means that the path traverses segment 1 in the forward direction, segment 2 in the forward direction, and segment 3 in the reverse direction. Additionally, the number of centromeres present in each path is reported, which should ideally be one, indicating a valid chromosome structure.

### S2 Terminal Event Simulation and Validation

We simulated a total of 117 terminal Structural Variations. During analyses, it was found that only 50 of the 117 SVs (42.7%) were reconstructed under the 0.2 Mbp matching distance (same distance used for non-terminal SVs; Supplementary Table S1). However, 78 of the 117 SVs (66.7%) were reconstructed under the 5 Mbp matching distance. Because one of the endpoint was in the terminal masking region, most terminal SV edge did not have an SV call. For unbalanced SVs, OMKar reconstructed these variations using the information from CNV calls, which had much greater error in the event boundaries. For inversion, as a balanced SV, missing the SV call meant OMKar had no other information on the SV, thus, resulting in a much lower recall rate at all distances. In addition, terminal events such as arm deletion may result in a false loss of centromere call, which result in a relatively higher false positive chromosomal-loss aneuploidy reconstruction from OMKar.

From the real data we received, we observed no terminal structural variation, hinting that terminal SVs were far less frequent than our simulation. For future reference, OMKar may incorporate additional information such as alignment contigs to improve the boundary accuracy and recall rates.

### S3 OMKar details

#### S3.1 Filtering SV and CNV calls

OMKar utilizes the CNV and SV calls from the Bionano pipeline. We applied several filters to improve data quality. First, we filtered out low-confidence CNV calls, defined as those with a confidence level of 0.95 or below, as well as those located in masked regions that could interfere with the analysis. Next, we excluded CNVs smaller than 200 kbp. However, if an excluded CNV was supported by a corresponding SV, it was retrieved during later processing, ensuring no relevant variations were missed.

Following these steps, SV calls were filtered based on variation-specific confidence thresholds established by BioNano pipeline: translocations (*T*_*trans*_ = 0.05), inversions (*T*_*inv*_ = 0.7), and indels (*T*_*indel*_ = 0). Additionally, SV calls in masked regions were discarded. Breakpoints from SVs were processed by sorting them based on chromosomal and genomic coordinates and merging adjacent breakpoints within a 50 kbp window to simplify the breakpoint graph. To ensure accurate representation, CNV segments were split if breakpoints occurred within their boundaries, guaranteeing that all breakpoints exclusively connect the terminal coordinates of segments.

#### S3.2 Converting to linear constraints

To tackle the inherent non-linearity of the absolute value and sign functions within the optimization problem, we introduce new variables and constraints to linearize these functions.

To convert the sign function into linear constraints, we employed the following approach: The ILP formulation for 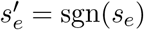 when *s*_*e*_ ≥ 0 is:

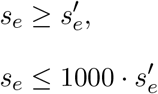

where 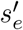 is a binary variable. This formulation ensures that 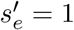 when *s*_*e*_ *>* 0 and 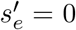 when *s*_*e*_ = 0, while maintaining linearity in the constraints. The constant 1000 is used as a sufficiently large value to approximate an upper bound on *s*_*e*_.

The absolute value function can be linearized *y*_*v*_ = |*x*_*v*_| as follows:

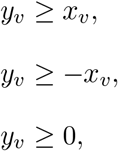

where *y*_*v*_ represents the absolute value of *x*_*v*_. These constraints ensure that *y*_*v*_ is equal to |*x*_*v*_| by considering both the positive and negative cases for *x*_*v*_. Since our objective is to minimize *y*_*v*_, it naturally corresponds to |*x*_*v*_|. Thus:

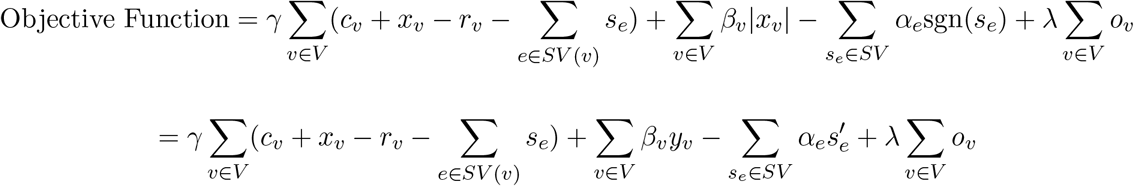

#### S3.3 Parameter determination

In determining *α*_*e*_, we consider the characteristics of structural variation *e*. If *e* signifies a high-confidence call and connects two distinct chromosomes, we set *α*_*e*_ = 72 otherwise, *α*_*e*_ = 9. A higher value is given to intra-chromosomal translocations to prioritize their preservation in the final karyotype. For the calculation of *β*_*v*_, we introduce a penalty parameter that becomes more pronounced as segments grow in length. To achieve this, we define 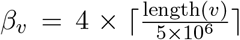. This parameter *β*_*v*_ ensures that the penalty for modifying longer segments is appropriately weighted.

The parameter *γ* is empirically determined and set to a value of 5, while *λ* is fixed at 1 to ensure that if two candidate decompositions are nearly identical, our preference is to select the one with fewer odd vertices. This empirical choice addresses the unique requirements of our approach, providing a balanced framework for the optimization process (Supplementary Table S4).

#### S3.4 OMKar Eulerian Path algorithm

To reconstruct chromosomal structures from breakpoint graphs, OMKar computes an Eulerian path, ensuring each edge is traversed exactly once while maintaining biologically meaningful constraints. It begins with a breakpoint graph, where vertices represent segment boundaries, and edges denote segment continuity, reference adjacencies, or breakpoint rearrangements. To enforce chromosomal structure, the algorithm prioritizes reference edges (minimizing deviation from the reference genome), followed by breakpoint edges (capturing structural variations), and lastly, segment edges. Using a recursive depth-first traversal, it starts from a telomeric vertex—representing a natural chromosomal endpoint—and evaluates edges with a validity function that prevents breaking segment integrity or disconnecting essential graph components. The pseudo-code is given in Algorithm 1.

##### Algorithm 1

Find Eulerian Tour Starting from Vertex *v* in Graph *G*

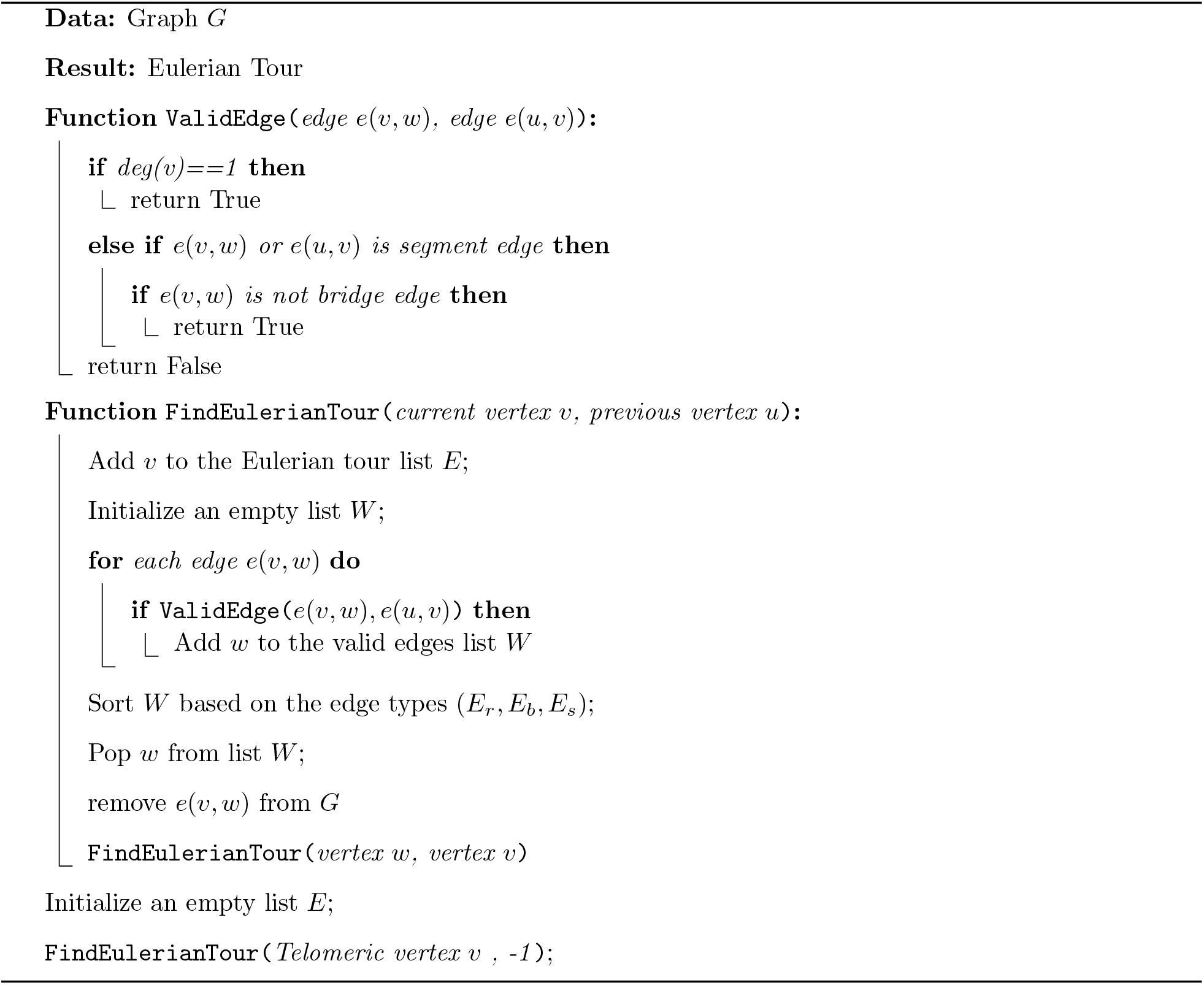

#### S3.5 OMKar Report: Event Interpretation

After segregating the chromosomes, OMKar interprets the structural variations in each chromosomal cluster using ISCN notation(Hastings et al., 2024) as follows:

1. For each reconstructed chromosome in the chromosomal cluster, its chromosomal identity is assigned based on the highest represented centromere, and if it is acentric, by the overall highest-represented chromosome in the remaining segments.
2. In a pre-processing step, the Wild Type (WT) chromosome corresponding to the reconstructed chromosome is segmented prior to alignment, using the segments in the reconstruction.
3. Next, OMKar performs an alignment between each reconstructed chromosome and the corresponding Wild Type (WT) using an alignment that maximizes the longest common subsequence, with a linear penalty for indels.
4. After alignment, blocks of aligned segments are separated into three types: 1) concordant block represents matching between the reconstruction and the WT, 2) insertion block represents segments inserted in the reconstruction, and 3) deletion block represents segments deleted in the reconstruction. Adjacent blocks of the same type, and representing contiguous genomic coordinates are merged to minimize the total number of blocks.
5. The final interpretation assigns a representative SV name (using ISCN nomenclature) to each insertion or deletion block, reporting a potential cause of the deviation from the WT. Each SV has its unique signature in the combination of block types (Supplementary Table S5). For example, an inversion is always an insertion block of inverted segments next to a deletion block of non-inverted segments. In this example, it is more likely that a single inversion resulted in both the deletion and the insertion, instead of two separate events (inverted insertion and deletion). Similarly during the interpretation step, a compound SV is preferred over a sequence of simpler SVs (preference is given to reducing the total number of events).

To minimize the number of events, event types are resolved from the most complex to the least, using the following preference order: (1) inter-chromosomal balanced translocation and transposition; (2) intra-chromosomal balanced translocation and transposition; (3) other intra-chromosomal variations. Finally, if any deletions or insertions remain unaccounted for, they are marked as simple deletions or duplicated-insertions. During each variation type’s resolution, each un-resolved block is iterated over, with the goal of being associated with the signature of that variation type. Balanced translocations attempt to associate anywhere in the cluster or chromosome, for inter- and intrachromosomal search, respectively. All other variations associate adjacent blocks.

For balanced translocations, the following step is performed after all insertion and deletion blocks are resolved. All balanced translocations are initially denoted as *transposition*, associating a deletion block and a non-adjacent insertion block of an overlapping set of segments, with allowance for small indels. When transpositions form a cycle, they are interpreted as *balanced translocations*. This is implemented recursively, by jumping between each associated deletion-insertion pair, and then looking for nearby transposition blocks of the opposite type. When exhausted, if a cycle is formed, an *n*-break balanced translocation is interpreted, and otherwise, all transpositions are interpreted as individual transpositions.

When associating between different blocks, a procedure called “seed-matching” is applied. For each block that is currently being resolved, it searches for an associated block, such that the common subsequence between the blocks (in indivisible-unit of each segment) is sufficiently long (10 kbp by default). The sizes of the flanking segments not matched may be limited by an “indel allowance”. For example, an insertion block of (*B*−, *A*−) next to a concordant block of (*B* + *C*+) will be interpreted as a left-duplication-inversion only if *A*− is less than 50 kbp. On the other hand, the size of *C*+ is irrelevant for associating a duplication-inversion, as it is not between the two blocks. These allowances are determined empirically (Supplementary Section S4).

#### S3.6 OMKar Report: Visualization

The visualization is achieved by intersecting the coordinates in the observed chromosomal segment list with a given cytoband coordinate table. Each band pattern is then stacked and displayed using Matplotlib (v3.7.5). In an alternative plot, the bands are segregated by the segment boundaries in the OMKar output, instead of the cytoband. The event label is then applied to the corresponding segment location on the band stack.

By default, the table of important genes comes from genes that have CN alteration or within 20 kbp of an event’s breakpoint. This list of genes are further filtered to only include those in the Developmental Disorders panel in the Gene2Phenotype database (DDG2P)(Thormann et al., 2019).

### S4 Analyses of Event Distances

The resolution of OGM was determined by the matching distances between the Truth SV edge and the reconstructed SV edge, using KarCheck. Observations from this analyses were used to estimate the sensitivity of OMKar event interpretation (Supplementary Table S5).

When applying KarCheck between the simulated and the reconstructed karyotypes, history logs from KarSim’s output was used to label SV edges on *K*_*t*_. This step marks each SV edge with its causal rearrangement. During KarCheck matching, the distance between a pair of matched edges were recorded with the causal SV type. This distance is further separated as the two distances of the endpoint.

Then, the matched edge from *K*_*t*_ was searched in OGM’s SMAP output (contains all SV calls), with a proximity of 50 kbp, equal to the SV call merging distance of OMKar. From Fig. S7, we observed duplication inversions tend to have much larger distances than all remaining structural variations simulated. Most of the non-duplication-inversion were reconstructed with less than 5 kbp distance from the truth, much lower than the average gene size of 10-15 kbp. Thus, if an SV contains or interrupts a gene, OGM and OMKar are likely to reconstruct the correct boundary to perform genotype-to-phenotype inference. Additionally, for all distances greater than 50 kbp, the true SV edge was not captured with a high proximity in the SMAP, therefore, OMKar either had to reconstruct based on a distant SV call or had to infer the missing SV call based on CNV call.

### S5 KarSim

#### S5.1 KarSim module for simulating karyotypes

The KarSim module outputs a molecular karyotype file, a corresponding FASTA file, and a history log with event segments and edges noted are outputted for downstream usage. The molecular karyotype and history log can be used for KarCheck comparison, while the FASTA file can be used as input for simulating the sequencing technology of choice, given that many of such simulators are already available. KarSim is publicly available at https://github.com/MolecularKaryotype/KarSimulator.

Three steps are taken in order, with step 2 being optional:

1. A template karyotype is created given the counts of autosomes and sex chromosomes.
2. (Optional) a series of SVs are applied to the karyotype
3. The FASTA formatted sequence of the rearranged chromosomes is generated.

All intermediate files in steps 1 and 2 are molecular karyotypes, which can be read-in for multiple parallel edits or outputting the FASTA file.

There are two methods to introduce additional SVs to a karyotype: 1) manual addition of SVs given the variation type and exact boundaries, and 2) using a parameter JSON file that contains the number of SVs, each SV type’s likelihood, max/min size for each SV type, terminal occurrence likelihood etc. Both processes can be applied multiple times and in an interleaved fashion. The types of SVs supported can be found in Supplementary Table S6. In addition, the user has the option to include a masking file such that SVs are not generated within proximity to any of these regions. Another parameter can be passed in to prevent events that result in the formation of segments smaller than a certain size, useful for sequencing technologies which have a resolution limit.

### S5.2 Implementation

KarSim is implemented using Python (version 3.9). A flowchart of the algorithm is in Supplementary Fig. S8A. Each chromosome in a karyotype is represented as a sequence of oriented segment objects of the hg38 genome, denoted by the chromosome, start index, and end index. To simulate an SV, left and right breakpoints are first introduced to the chromosome, breaking up the chromosome into segments. This results in each SV breakpoint being on the exact border of a segment. Then, the corresponding rearrangement is applied to the segments to simulate the intended SV.

For the parameterized-random SV selection, SVs are selected one at a time, for the number of SVs indicated on the parameter file as follows:

1. The SV type is randomly selectedbased on its likelihood.
2. The event size is selected from a uniform distribution between the maximum and minimum size indicated for the SV type.
3. The left breakpoint of the event location is selected uniformly among all chromosomes.
4. The right breakpoint is calculated based on the size of the event selected earlier. For a balanced reciprocal translocation, it is selected similar to the left breakpoint.
5. The breakpoints are applied to partition the segments, and if a masked region file or a smallest segment allowance is applied, the resulting breakpoints and segments are checked for legality. If the result is illegal under the parameters, steps three through five are recomputed.

#### S5.3 Generation and Processing of Simulated OGM Data

The FASTA files generated by the KarSim module were processed by OMSim(Miclotte et al., 2017) to simulate a BNX file containing OGM molecules with added noise. Parameters for the enzyme, OGM generation, and noise were sourced from publicly available repositories(RaeisiDehkordi, 2024). The Bionano Solve pipeline (v3.7)(BionanoGenomics, 2018) was then used to compute CNVs, SV calls, and contig alignments, and these outputs were used as input for OMKar to reconstruct the final virtual karyotype.

When simulating structural variations, we used the masking files provided in Bionano Solve v3.7 (also included in repository (RaeisiDehkordi, 2024)). The masking file is on reference hg38, with size of 423.1 Mbp (13.65%), including 128.1 Mbp (4.13%) in centromeres and telomeres, and 64.5 Mbp (2.15%) in acrocentric chromosomes’ p-arm. None of the SVs was simulated with breakpoint boundary within 200 kbp of any masking region. Events were simulated with size 50 kbp to 2 Mbp.

### S6 KarCheck

KarCheck is designed to be a symmetric comparator between two unphased karyotypes, denoted by *K*_*r*_ and *K*_*t*_, by comparing their SV and CNV calls. In addition, to accommodate molecular techniques that do not have a nucleotide-level resolution, KarCheck has an adjustable tolerance for small breakpoint mis-matching for an SV that is present in both karyotypes. KarCheck is publicly available at https://github.com/MolecularKaryotype/KarComparator.

#### S6.1 KarCheck Preprocessing

Preprocessing is first applied to *K*_*t*_ and *K*_*r*_ to partition chromosome groups into *chromosome clusters*. Two chromosome groups are linked into the same cluster when there exists a breakpoint connecting them (signaling an inter-chromosomal SV). A maximal connected component of linked chromosome groups is denoted as a chromosome cluster.

Recall that we define each karyotype as an ordered list of segments. To make the two karyotypes comparable, we further partition their segments such that the two karyotypes share an identical set of segments (Supplementary Fig. S8B). To achieve this, all left/right endpoints of segments are collected, and if a segment has an endpoint internal to it, it is split into two, ensuring the left/right endpoint are on the boundaries (Supplementary Fig. S8B).

#### S6.2 SV similarity computation

After pre-processing, the SVs between the two karyotypes are compared as a directed-multi-graph, which offers ease of checking the orientation of each SV edge (Supplementary Fig. S8C). On this graph, nodes represent either the start or the end of a segment (denoted as s and t), with the addition of a source node (*S*) and a sink node (*T*) for transition at the start and end of each chromosome. Edges represent the presence of a transition between two segments. For example, a chromosome of [*k*+, *l*-], where the WT is [*k*+, *l*+], will be represented by edges (*S, k*_*s*_),(*k*_*s*_, *l*_*t*_), and (*l*_*s*_, *T*).

Since the two karyotypes share the same set of segments, they also share the same set of nodes. Therefore, the comparison between the two sets of edges on this graph is equivalent to the comparison of the two sets of SVs. To allow tolerance in breakpoint matching, we define a linear distance function. Denote two non-segment edges as (*a, b, o*) and (*c, d, o*^*′*^), where *o* (and *o*^*′*^) describe orientation. Define distance *D*((*a, b, o*), (*c, d, o*^*′*^)) as:

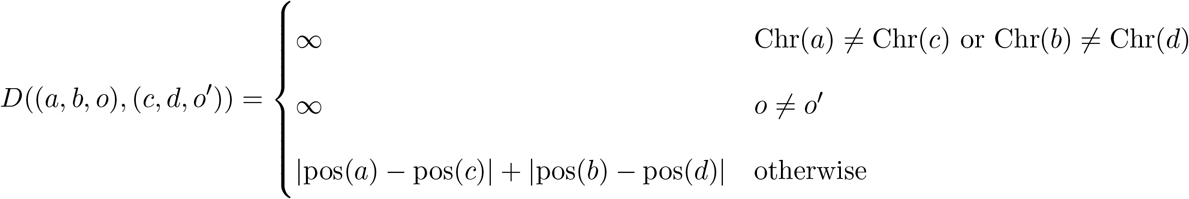

Prior to distance computation, identical edges between the two graphs are pruned. Second, if a transition is s-to-t or t-to-s and is between two segments from the same chromosome with a small distance (≤ 5 kbp), it is pruned. This is justified by that these transitions represent small indels without change in orientation, which are not responsible if the technique has a minimum resolution threshold. Finally, minimum weight bipartite matching is performed for the remaining transition edges between the two karyotypes, with a maximum allowed matching distance 200 kbp. Matching pairs are pruned, because they are considered similar SV edges. The matching distances are recorded for downstream analyses upon resolution. Finally, all residual non-segment edges are the differential SV edges between the two karyotypes. We use *K*_*r*_ to denote the reconstructed chromosome, and *K*_*t*_ as the WT or true chromosome. Therefore, residual edges in *K*_*t*_ represent false negatives, and residual edges in *K*_*r*_ represent false positives.

##### SV Similarity Metrics

The similarity of each individual SV edge is compared via the count of *K*_*t*_ edges after the two initial pruning procedures, and the residual edges in *K*_*t*_ and *K*_*r*_ after final matching. TP = |inital edge| − |*K*_*t*_ residual|, FN = |*K*_*t*_ residual|, and FP = |*K*_*r*_ residual|. A Jaccard Similarity is computed for each cluster of compared simulated data to penalize both false positive and false negative events. For comparison against real data, only the recall is computed for each cluster, as the cytogenetic methods employed for the reference calls do not necessarily have small enough resolution or ability to catch de-novo balanced events.

#### S6.3 Copy Number similarity comparison and metrics

CN comparison is done by binning the whole genome (excluding prefix/suffix masked region) into spanning, non-intersecting bins of 50 kbp +/-100 bp (exact size chosen to maximize the size of the last bin on the chromosome). Each bin is used to store the average CN within that region, separately for the *K*_*t*_ and the *K*_*r*_.

Then, for each cluster, the chromosome groups are determined by the union of all the chromosomal origins of the segments in the cluster. Each cluster’s CN bins are the subset of the total CN bins, to only include the chromosome groups within the cluster. A WT expected count is determined to each chromosome group by rounding the average bin CN from *K*_*t*_ (diploid for autosomes and XX or XY for sex chromosomes).

The values of the CN bins’ of *K*_*t*_ and *K*_*r*_ are computed with CNs from all corresponding segments. If a bin has a gain or loss of more than 0.05 CN from the WT expected count, it is a marked as “CN gain” or “CN loss”. Otherwise, it is marked as “CN neutral”. This forms a paired CNV array where the Jaccard Similarity can be computed. The denominator of the similarity is the count of bins marked as CN gain or loss in either *K*_*t*_ or *K*_*r*_, and the numerator is the count of bins where *K*_*t*_ and *K*_*r*_ agree on CN gain or loss.

#### S6.4 Addtional Functionalities for Downstream Analyses

Edges on both *K*_*t*_ and *K*_*r*_ can be labeled with additional input. For example, Table 2 was computed where each edge in *K*_*t*_ from the simulation was labeled with the Structural Variation event type, so a summary statistics was generated from the residual edge count of each event type.

When matching transition edges, the distances between the matched edges can be collected for analysis. This can be further categorized by having *K*_*t*_’s transition edge labeled with the causal event type. A detailed analysis on OMKar’s reconstructions’ distances against the simulation can be found in Supplementary Section S4.

#### S6.5 Usage in validating real data reconstructions with previous cytogenetic records

A function was implemented in KarCheck to allow efficient input of a karyotype for the purpose of validating a reconstruction against previous cytogenetic records on that karyotype. For each karyotype, its aneuploidy (if any), each event’s induced SV-breakpoints, and CN changes are taken as input for the validation. This information is sufficient to populate a full molecular karyotype, while KarCheck assumes the rest of the genome is WT.

For real data with cytogenetic records, the cytogenetic records were used as the “truth” karyptype (*K*_*t*_) and compared against the reconstruction (*K*_*r*_). CN gains called with microarray do not specify the structural breakpoints of the amplification. For these, we first assumed each amplification was a tandem duplication, and if the matching failed, we manually verified if the reconstruction contained the amplification as a segmental duplication. Additionally, previous cytogenetic techniques using microarray and staining did not fully capture every event on the genome, so the *K*_*t*_ was treated as incomplete, with potentially missed TP events. Therefore, for our final statistics of the comparison, we only computed the recall to verify if OMKar reconstruction included all the previously identified TPs.

